# GWAS meta-analysis of psoriasis identifies new susceptibility alleles impacting disease mechanisms and therapeutic targets

**DOI:** 10.1101/2023.10.04.23296543

**Authors:** Nick Dand, Philip E Stuart, John Bowes, David Ellinghaus, Joanne Nititham, Jake R Saklatvala, Maris Teder-Laving, Laurent F Thomas, Tanel Traks, Steffen Uebe, Gunter Assmann, David Baudry, Frank Behrens, Allison C Billi, Matthew A Brown, Harald Burkhardt, Francesca Capon, Raymond Chung, Charles J Curtis, Michael Duckworth, Eva Ellinghaus, Oliver FitzGerald, Sascha Gerdes, Christopher E M Griffiths, Susanne Gulliver, Philip Helliwell, Pauline Ho, Per Hoffmann, Oddgeir L Holmen, Zhi-ming Huang, Kristian Hveem, Deepak Jadon, Michaela Köhm, Cornelia Kraus, Céline Lamacchia, Sang Hyuck Lee, Feiyang Ma, Satveer K Mahil, Neil McHugh, Ross McManus, Ellen H Modalsli, Michael J Nissen, Markus Nöthen, Vinzenz Oji, Jorge R Oksenberg, Matthew T Patrick, Bethany E Perez-White, Andreas Ramming, Jürgen Rech, Cheryl Rosen, Mrinal K Sarkar, Georg Schett, Börge Schmidt, Trilokraj Tejasvi, Heiko Traupe, John J Voorhees, Eike Matthias Wacker, Richard B Warren, Rachael Wasikowski, Stephan Weidinger, Xiaoquan Wen, Zhaolin Zhang, BSTOP study group, Estonian Biobank research team, Anne Barton, Vinod Chandran, Tõnu Esko, John Foerster, Andre Franke, Dafna D Gladman, Johann E Gudjonsson, Wayne Gulliver, Ulrike Hüffmeier, Külli Kingo, Sulev Kõks, Wilson Liao, Mari Løset, Reedik Mägi, Rajan P Nair, Proton Rahman, André Reis, Catherine H Smith, Paola Di Meglio, Jonathan N Barker, Lam C Tsoi, Michael A Simpson, James T Elder

**Affiliations:** Department of Medical & Molecular Genetics, School of Basic & Medical Biosciences, Faculty of Life Sciences & Medicine, King’s College London, London, UK; Health Data Research UK, London, UK; Department of Dermatology, University of Michigan Medical School, Ann Arbor, MI, USA; Centre for Genetics and Genomics Versus Arthritis, The University of Manchester, Manchester, UK; National Institute for Health and Care Research (NIHR) Manchester Biomedical Research Centre, The University of Manchester, Manchester, UK; Institute of Clinical Molecular Biology, Christian-Albrechts-University of Kiel, Kiel, Germany; Deparment of Dermatology, University of California San Francisco, San Francisco, CA, USA; Institute of Genomics, University of Tartu, Tartu, Estonia; Department of Clinical and Molecular Medicine, NTNU - Norwegian University of Science and Technology, Trondheim, Norway; K.G. Jebsen Center for Genetic Epidemiology, Department of Public Health and Nursing, NTNU - Norwegian University of Science and Technology, Trondheim, Norway; BioCore - Bioinformatics Core Facility, NTNU - Norwegian University of Science and Technology, Trondheim, Norway; Clinic of Laboratory Medicine, St.Olavs Hospital, Trondheim University Hospital, Trondheim, Norway; Department of Dermatology and Venereology, Institute of Clinical Medicine, University of Tartu, Tartu, Estonia; Institute of Human Genetics, Universitätsklinikum Erlangen, FAU Erlangen-Nürnberg, Erlangen, Germany; RUB University Hospital JWK Minden, Department of Rheumatology, Minden, Germany; Jose-Carreras Centrum for Immuno- and Gene Therapy, University of Saarland Medical School, Homburg, Germany; St John’s Institute of Dermatology, School of Basic & Medical Biosciences, Faculty of Life Sciences & Medicine, King’s College London, London, UK; Division of Translational Rheumatology, Immunology - Inflammation Medicine, University Hospital, Goethe University, Frankfurt am Main, Germany; Fraunhofer Institute for Translational Medicine and Pharmacology ITMP, Frankfurt am Main, Germany; Fraunhofer Cluster of Excellence Immune-mediated Diseases CIMD, Frankfurt am Main, Germany; Division of Rheumatology, University Hospital, Goethe University, Frankfurt am Main, Germany; Genomics England, Canary Wharf, London, UK; Institute of Psychiatry, Psychology and Neuroscience, King’s College London, Denmark Hill, Camberwell, London, UK; National Institute for Health and Care Research (NIHR) Biomedical Research Centre, South London and Maudsley Hospital, London, UK; UCD School of Medicine and Medical Sciences and Conway Institute of Biomolecular and Biomedical Research, University College Dublin, Ireland; Department of Dermatology, Venereology and Allergy, University Hospital Schleswig-Holstein, Campus Kiel, Kiel, Germany; Centre for Dermatology Research, University of Manchester, NIHR Manchester Biomedical Research Centre, Manchester, UK; Department of Dermatology, King’s College Hospital NHS Foundation Trust, London, UK; Newlab Clinical Research Inc, St. John’s, NL, Canada; National Institute for Health and Care Research (NIHR) Leeds Biomedical Research Centre, Leeds Teaching Hospitals Trust, UK; Leeds Institute of Rheumatic and Musculoskeletal Disease, University of Leeds, UK; The Kellgren Centre for Rheumatology, Manchester University NHS Foundation Trust, Manchester, UK; Institute of Human Genetics, University of Bonn, School of Medicine & University Hospital Bonn, Bonn, Germany; HUNT Research Centre, Department of Public Health and Nursing, NTNU - Norwegian University of Science and Technology, Levanger, Norway; Levanger Hospital, Nord-Trøndelag Hospital Trust, Levanger, Norway; Department of Medicine, University of Cambridge, Cambridge, UK; Division of Rheumatology, Geneva University Hospital, Geneva, Switzerland; Department of Computational Medicine and Bioinformatics, University of Michigan, Ann Arbor, MI, USA; St John’s Institute of Dermatology, Guy’s and St Thomas’ National Health Service (NHS) Foundation Trust, London, UK; Royal National Hospital for Rheumatic Diseases and Dept Pharmacy and Pharmacology, University of Bath, UK; Department of Clinical Medicine, Trinity Translational Medicine Institute, Trinity College Dublin, Ireland; Department of Dermatology, Clinic of Orthopedy, Rheumatology and Dermatology, St. Olavs Hospital, Trondheim University Hospital, Trondheim, Norway; Department of Dermatology, University of Münster, Münster, Germany; Weill Institute for Neurosciences, Department of Neurology, University of California, San Francisco, CA, USA; Department of Dermatology, Northwestern University, Evanston, IL, USA; Department of Internal Medicine 3, Friedrich-Alexander-University Erlangen-Nürnberg (FAU) and Universitätsklinikum Erlangen, Ulmenweg 18, 91054, Erlangen, Germany; Deutsches Zentrum Immuntherapie (DZI), Friedrich-Alexander-University Erlangen-Nürnberg and Universitätsklinikum Erlangen, Erlangen, Germany; Division of Dermatology, Toronto Western Hospital, University of Toronto, Toronto, Ontario, Canada; Institute of Medical Informatics, Biometry and Epidemiology, University Hospital Essen, University of Duisburg-Essen, Essen, Germany; Ann Arbor Veterans Affairs Hospital, Ann Arbor, MI, USA; Division of Musculoskeletal and Dermatological Sciences, School of Biological Sciences, Faculty of Biology, Medicine and Health, The University of Manchester, UK; Centre for Dermatology Research, Salford Royal Hospital, Northern Care Alliance NHS Foundation Trust, Manchester Academic Health Science Centre, Manchester, M6 8HD, UK; Department of Biostatistics, Center for Statistical Genetics, University of Michigan, Ann Arbor, MI, USA; Schroeder Arthritis Institute, Krembil Research Institute, and Toronto Western Hospital, University Health Network and University of Toronto, Toronto, Ontario, Canada; College of Medicine, Dentistry, and Nursing, University of Dundee, Dundee, UK; Department of Dermatology, Discipline of Medicine, Faculty of Medicine, Memorial University of Newfoundland, St. John’s, NL, Canada; Dermatology Clinic, Tartu University Hospital, Tartu, Estonia; Perron Institute for Neurological and Translational Science, Nedlands, WA 6009, Australia; Centre for Molecular Medicine and Innovative Therapeutics, Health Futures Institute, Murdoch University, Perth, WA 6150, Australia; Estonian Genome Centre, Institute of Genomics, University of Tartu, Tartu, Estonia; Memorial University of Newfoundland, St. John’s, NL, Canada

## Abstract

Psoriasis is a common, debilitating immune-mediated skin disease. Genetic studies have identified biological mechanisms of psoriasis risk, including those targeted by effective therapies. However, the genetic liability to psoriasis is not fully explained by variation at robustly identified risk loci. To move towards a saturation map of psoriasis susceptibility we meta-analysed 18 GWAS comprising 36,466 cases and 458,078 controls and identified 109 distinct psoriasis susceptibility loci, including 45 that have not been previously reported. These include susceptibility variants at loci in which the therapeutic targets IL17RA and AHR are encoded, and deleterious coding variants supporting potential new drug targets (including in *STAP2*, *CPVL* and *POU2F3*). We conducted a transcriptome-wide association study to identify regulatory effects of psoriasis susceptibility variants and cross-referenced these against single cell expression profiles in psoriasis-affected skin, highlighting roles for the transcriptional regulation of haematopoietic cell development and epigenetic modulation of interferon signalling in psoriasis pathobiology.

## Introduction

Psoriasis is a common immune-mediated skin disease with significant impact on psychosocial wellbeing, lifelong morbidity, and mortality.^1, 2^ With an estimated 60 million people affected worldwide,^3^ it represents a substantial economic burden.^4, 5^

Psoriasis has a strong genetic component, with heritability estimated at 66%.^6^ Previous genome-wide association study (GWAS) meta-analyses have identified 65 genomic loci at which genetic variation is associated with psoriasis susceptibility in European ancestry populations,^7–9^ and 17 more reported in Asian ancestry populations.^10^ A substantial fraction of genetic risk is attributed to the Major Histocompatibility Complex (MHC) class I allele *HLA-C*06:02* and related antigen processing and presentation functions, while pathogenic roles for the IL-23/IL-17 immune axis, type I interferons and NF-κB have also been established.^7, 11–13^ Evidence that genetic variation influences psoriasis risk through these pathways underscores the remarkable consistency between genetic perturbations and the effectiveness of biologic therapies targeting IL-23 and IL-17.^1^

While they have greatly influenced current models of psoriasis pathobiology, previous GWAS meta-analyses of psoriasis have been modest in size compared to those of other common diseases in the current era of population-based bioresources.^14^ Larger studies offer enhanced ability to detect genetic associations with small effects and discriminate independent association signals within loci. There remain open questions around how, and in which cell types, the presence of risk-increasing alleles can lead to the dysregulated immune processes that characterise psoriasis. Unpacking these causal mechanisms and better understanding their heterogeneity across individuals should help inform how existing targeted therapies can be deployed to specifically disrupt the inflammatory loop underlying psoriasis while limiting unintended adverse consequences,^15, 16^ and suggest new therapeutic targets for patients in whom current treatments are ineffective or response is transient.^17^ It will also begin to explain the mechanistic basis for the high burden of co-morbidities suffered by individuals with psoriasis.^18, 19^

Here we report a meta-analysis of 18 case-control genome-wide association studies conducted by an international consortium to increase the statistical power for genetic discovery in psoriasis, and to characterise causal variants, genes, pathways, and cell types.

## Results

### Discovery of new psoriasis susceptibility regions

To identify genomic loci at which genetic variation is associated with psoriasis susceptibility, we performed fixed-effect standard-error-weighted GWAS meta-analysis for a total of 11,808,957 autosomal variants across 18 studies comprising a total of 494,544 unrelated European ancestry individuals. The genomic inflation factor^20^ (λ_GC_) of 1.14 and LD score regression intercept of 1.07 indicate modest inflation of the meta-analysis test statistics that is partially driven by polygenicity, consistent with other complex diseases.^21^

Consistent with previous GWAS of psoriasis, by far the strongest evidence of association was observed within the MHC region on chromosome 6. The association signal peaks at rs12189871 (OR: 3.31, 95% CI 3.21-3.40, P=1.8×10^-1524^) but genome-wide significant evidence of association (P<5×10^−8^) was observed from positions chr6:25,622,875 telomeric to and chr6:33,971,609 centromeric to the MHC, reflecting multiple independent associations and complex patterns of linkage disequilibrium (LD)^22^ (Supplementary Figure 1).

Genome-wide significant psoriasis associations were also observed in the present study at all but two of the previously reported susceptibility loci in Europeans^7–9^ (Figure 1, Supplementary Table 1). The two loci without genome-wide significant evidence of association (13q14.11 and 21q22.12) were previously reported as psoriasis susceptibility loci in a trans-ethnic meta-analysis^23^ and fell just short of genome-wide significance in the current study (Supplementary Table 2).

**Figure 1.**
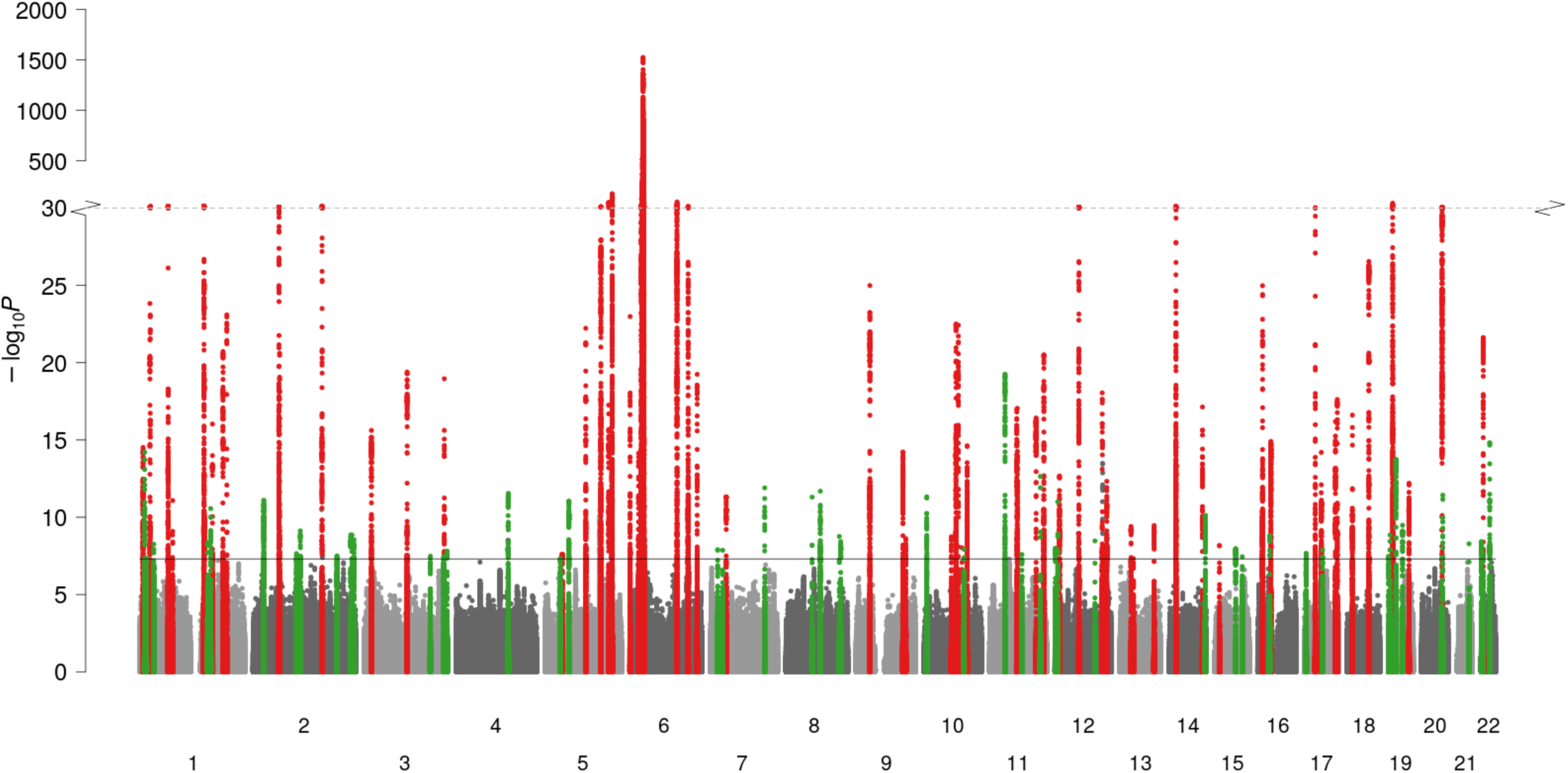
Manhattan plot summarising genome-wide associations with psoriasis susceptibility. x-axis, genomic position; y-axis, -log_10_(P-value) of association; red and green points, regions previously and newly associated, respectively, with psoriasis susceptibility at genome-wide significance (P=5×10^−8^) in European ancestry populations; solid horizontal line, genome-wide significance threshold; dotted horizontal line, y-axis break at 10^−30^; chromosomes (labelled 1-22) are alternately shaded for clarity.

We identified associated genetic variants at 109 distinct loci, 50 of which have not been previously implicated in psoriasis susceptibility in European ancestry populations (Figure 1, Supplementary Tables 1 and 3, Supplementary Note). Five of these 50 loci encompass variants previously reported at genome-wide significance in other ancestral populations.^22,24, 25^

To resolve the presence of multiple independent association signals at each of these loci we performed conditional and joint analysis^26^ across 108 susceptibility loci (excluding the MHC). We found evidence of two or more high-confidence independent associations at 27 loci, including evidence of two independent signals each at newly reported loci at 4q27 and 8q12.2. This totals 148 independent non-MHC psoriasis association signals (Supplementary Table 1).

Assuming a prevalence of 1.5%, we estimate that 46.5% (standard error, SE: 4.4%) of variance in the liability to psoriasis is explained by common SNPs outside of the MHC region, increasing to 59.2% (SE: 13.2%) when including the MHC region. The 52 independent association signals at the 50 newly identified susceptibility loci were estimated to contribute 2.9% of the non-MHC common variant liability in addition to the 11.6% accounted for by established loci (Supplementary Table 4).

### Fine-mapping of candidate causal variants

Within each susceptibility locus, we sought to identify variants with strong statistical and functional evidence of being the causal variant underlying the psoriasis association signal. We constructed Bayesian 95% credible sets for causal variants at 144 sufficiently well-imputed independent susceptibility signals (Methods; Supplementary Table 1). A single variant had a posterior probability of >0.5 for being causal (PP_max_>0.5) at 51 (36%) of the fine-mapped non-MHC signals with resolution to a single putative causal variant (PP_max_>0.95) for 22 signals (15%) (Supplementary Table 5).

Our expanded GWAS meta-analysis provides increased power and improved ability to resolve causal variants. Thus, comparison against fine-mapping in the previous meta-analysis^7^ revealed the same or fewer number of variants in 95% credible sets at 52 (83%) of the 63 established psoriasis susceptibility loci (Figure 2A, Supplementary Figure 2, Supplementary Table 6).

**Figure 2.**
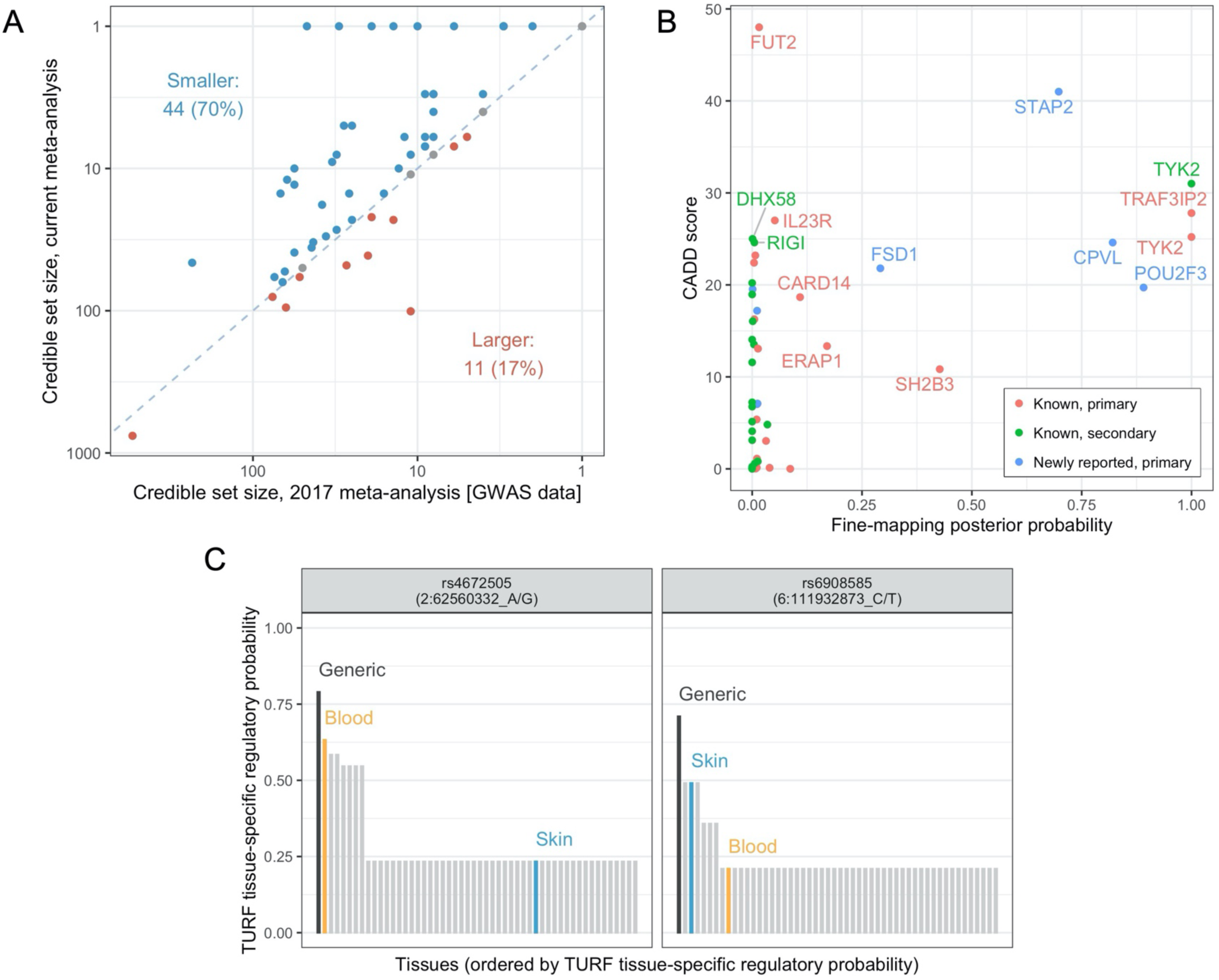
Statistical and functional fine-mapping. **A**. Comparison of 95% Bayesian credible sets to previous GWAS meta-analysis. Each point represents a different association signal established in the previous meta-analysis (Tsoi et al., 2017). Point colour indicates direction of change, blue dashed line indicates equality. **B**. Prioritisation of protein-altering variants. Points represent protein-altering variants identified in Bayesian credible sets for independent psoriasis signals; x-axis: posterior probability of causality from statistical fine-mapping analysis, y-axis: CADD score estimating deleteriousness of protein altering variant, point colour: whether corresponding susceptibility signal is in a known or newly reported genomic region and whether primary or secondary signal. Note the *TRAF3IP2* variant is rs33980500, discussed in the main text. **C**. Highlighted high-confidence regulatory variants derived from TURF analysis. For each variant, bars show the generic and tissue-specific regulatory probabilities (y-axis) estimated by TURF for all tissues (x-axis). Blood and skin are highlighted in orange and blue, respectively.

To further highlight likely disease-causing variants and identify the biological mechanisms through which they influence pathological processes, we functionally annotated 5,345 variants present in 95% credible sets. For 24 association signals, at least one variant in the credible set was predicted to give rise to altered or truncated protein sequences (Supplementary Table 7). Twenty of these missense and nonsense variants were predicted to be deleterious (CADD score > 15), including well-known alleles influencing psoriasis risk in *IL23R*, *TRAF3IP2*, *TYK2* and *RIGI* (formerly known as *DDX58*).^8^ Of relevance to *RIGI*, a deleterious missense variant was also observed in the structurally related gene *DHX58* (ENSP00000251642.3:p.Asn461Ser, CADD score 25.0), encoding the nucleic acid receptor LGP2 which, like the protein retinoic acid-inducible gene (RIG-I) encoded by *RIGI*, contributes to the innate antiviral response. A further six putative causal variants were observed in genes located at novel psoriasis susceptibility loci including high-confidence deleterious variants affecting *CPVL* (ENSP00000387164.1:p.Tyr168His) and *POU2F3* (ENSP00000260264.4:p.His154Arg), and a low-frequency nonsense allele in *STAP2* that is protective against psoriasis (ENSP00000468927.1:p.Tyr169Ter, CADD score 41.0, OR = 0.79, 95% CI 0.76-0.82) (Figure 2B).

We also assessed the likelihood that variants within the credible sets have a regulatory function, both across tissues and specifically within blood or skin.^27^ Consistent with the notion that statistical fine-mapping identifies causal variants, signals that resolve to smaller credible sets are enriched for variants with higher regulatory probability (P_generic_ = 2.0×10^-11^, P_tissue-specific_ = 1.8×10^-17^, Kruskal–Wallis test) (Supplementary Table 8, Supplementary Figure 3). At 14 association signals, a single candidate variant was prioritised with a high generic regulatory probability, both in absolute terms and relative to other variants in the credible set (Supplementary Table 9, Supplementary Figure 4). Notably, this included two independent signals at *TRAF3IP2*, a gene that encodes a regulator of NF-κB at chromosome 6q21.^28–30^ We found strong evidence that the well-known psoriasis-associated missense variant rs33980500 (p.Asp10Asn; Figure 2B) itself has a regulatory role, and independently that rs6908585 is a skin-specific regulatory variant (regulatory probability of 0.491; Figure 2C), suggesting that both coding and gene regulation of *TRAF3IP2* contribute to psoriasis susceptibility at this locus.

### Identification of candidate genes

Given the demonstrated importance of putative regulatory variants at many of the established and newly identified psoriasis susceptibility loci, we sought to identify genes whose expression is associated with psoriasis susceptibility variants in blood and skin (both sun exposed and unexposed) by means of a transcriptome-wide association study (TWAS) derived from Genotype-Tissue Expression (GTEx) project data.^31^ As expected given the large effects of psoriasis MHC associations and the extensive LD characteristic of the region, and as characterised in detail previously,^22^ there are multiple genes within the MHC with predicted expression differences (Supplementary Figure 5).

Outside of the MHC, transcriptome-wide significant gene expression differences (P<2.18×10^−6^; Methods) were observed at 47 susceptibility loci (12 newly reported) and a further six loci within 1Mb of association signals with suggestive evidence of association with psoriasis (P_meta_<10^-5^) (Supplementary Table 10). Interestingly, the gene predicted to be differentially expressed was the closest gene to the lead variant of the association signal at only 19 (40%) of these 47 loci (Supplementary Figure 6, Supplementary Table 11). Thirty-two of the non-MHC TWAS genes have been previously highlighted as candidate psoriasis genes.^10^ Amongst genes at new loci, *IRF5*, which encodes an interferon (IFN) regulatory factor that activates type I IFN responses, has been previously implicated in immune-mediated inflammatory disease through its association with systemic lupus erythematosus.^32, 33^

To identify groups of genes across psoriasis susceptibility loci with related biological function we employed DEPICT.^34^ This highlighted a series of immune pathways and gene sets whose membership is over-represented at psoriasis risk loci (1,632 gene sets at false discovery rate <5%; Supplementary Figure 7, Supplementary Table 12A); this includes protein-protein interaction subnetworks for the genes *CBL* (proto-oncogene), *EGFR* (epidermal growth factor receptor; EGFR signalling being regulated via CBL^35^), *ESX1* and *TEC* (Tec Protein Tyrosine Kinase) (Supplementary Table 12B) that are implicated for the first time in this study.

### Cellular and functional contexts of psoriasis associations

To identify the cellular contexts of genes whose transcription is mediated by psoriasis susceptibility variation we investigated expression patterns in single-cell transcriptomes derived from lesional and non-lesional skin of up to 14 chronic plaque psoriasis patients. 140 genes that were detectably expressed in scRNA-seq and identified in the TWAS analysis formed five clusters based on average expression patterns across twelve cell types (Figure 3). The clusters exhibited prominent upregulation of genes in: (1) lymphatic endothelial and eccrine cells, (2) keratinocytes, (3) T cells and dendritic cells, (4) melanocytes, and (5) fibroblasts, lymphatic endothelial cells and pericytes, respectively. As expected, the genes located in the epidermal differentiation complex at chr1q21.3 were preferentially expressed in epidermal keratinocytes. Genes involved in the antiviral response were identified as being enriched in the T cell/dendritic cell, melanocyte and keratinocyte clusters (Supplementary Figure 8).

**Figure 3.**
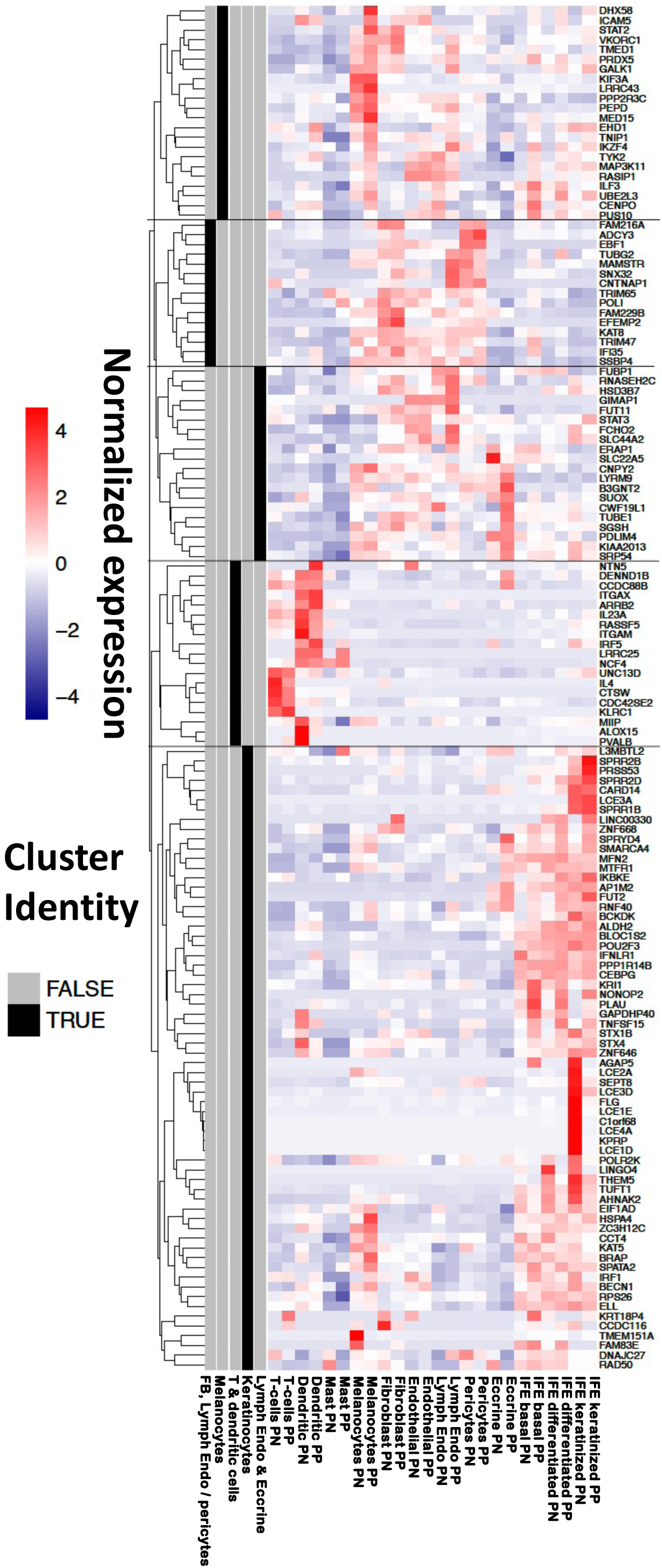
Relative expression of TWAS genes in single-cell skin transcriptomes of psoriasis patients. Expression level (cell colour; purple-red scale) represents mean value among cells in the corresponding cell type/condition (x-axis). For each gene (y-axis), the values were standardised and expression patterns were used for clustering (dendrogram, left hand side). Five clusters were identified (grey/black bars, left hand side) and labelled according to the cell types that exhibit highest expression for the genes in the cluster. PP, psoriasis lesions; PN, non-lesional skin; Lymph Endo, lymphatic endothelial cells; IFE, interfollicular epidermis. Clusters are identified based on enrichment for: (1) lymphatic endothelial and eccrine cells; (2) keratinocytes; (3) T cells and dendritic cells; (4) melanocytes; (5) fibroblasts, lymphatic endothelial cells and pericytes.

Despite the T cell centric regulation highlighted by previous studies, our results show that psoriasis-associated variants also govern gene regulation in stromal cells and keratinocytes. We therefore investigated whether specific cytokines regulate subsets of genes whose expression is influenced by psoriasis susceptibility variants, by examining the transcript abundance of non-MHC TWAS genes in transcriptome profiles from keratinocytes following a series of cytokine challenges (Supplementary Figure 9, Supplementary Table 13).^36^ The transcriptomic shift most strongly enriched for TWAS genes was induced by a combination of IL17A and TNF (FC=14.2; FDR=2.3×10^−5^), followed by TNF alone (FC=11.8; FDR=1.82×10^−4^).

These results suggest potential context-specific biological effects in keratinocytes for the implicated genes and highlight the roles of psoriasis loci in regulating inflammatory response in keratinocytes.

### Genome-wide correlations and causal inference

A series of recent studies have investigated the shared genetic architecture of psoriasis with other disease and health-related traits and have identified putative causal relationships with smoking, obesity and lifetime risk of cardiovascular disease.^37–40^ To further investigate the shared genetic liability and potential causal relationships, we assessed genetic correlation between psoriasis and 345 disease and health-related traits and evaluated asymmetry in the correlation structure (specifically, the mixed fourth moments between effect sizes^41^) that are consistent with causal relationships.

Positive genetic correlations were observed between psoriasis and 36 diseases and health-related traits (Supplementary Table 14) including evidence of a substantial shared genetic architecture with colitis, generalised pain, angina and pulmonary disorders (r_g_≥0.30, P<6.8×10^−6^ in all cases; Supplementary Figure 10). Significant genetic correlations were also observed with 42 physical and functional measures (Supplementary Figure 11), and 11 traits related to lifestyle and quality of life (Supplementary Figure 12).

Among traits for which we found evidence of a genetic correlation with psoriasis, 12 have evidence of asymmetry in the correlation structure that is consistent with a causal relationship (FDR<0.05) (Supplementary Table 15, Supplementary Figure 13). Susceptibility to stroke, triglyceride levels and multiple measures of adiposity have genetic support for a putative causal role in psoriasis (all genetic causal proportion (GCP)<-0.67, P≤4.6×10^−3^), the latter consistent with previous reports using Mendelian randomisation.^37, 38^ Notably, we also observed evidence indicating a causal role of psoriasis genetic risk in the development of other traits including back pain and generalised pain, fracture risk, diabetes and periodontitis (all GCP>0.63, P<3.2×10^−3^).

## Discussion

This work represents the largest genetic study of psoriasis susceptibility undertaken to date. Relative to previous efforts^7^, the increased statistical power of our larger meta-analysis sample size has resulted in the identification of 45 new psoriasis susceptibility loci with genome-wide significant evidence of association. A further five risk loci have been observed for the first time at genome-wide significance in populations of European ancestry.

A range of recent and emerging targeted therapies have proven highly effective in psoriasis.^1^ Many of these exemplify the concordance between a drug’s efficacy and the presence of disease susceptibility variants that disrupt the targeted pathway,^17^ including biologics targeting IL-23 (encoded by *IL23A* and *IL12B* at the 12q13.3 and 5q33.3 psoriasis susceptibility loci, respectively) and deucravacitinib^42^ a small-molecule TYK2 inhibitor whose development was directly informed by the presence of loss-of-function alleles of *TYK2* that confer protection to psoriasis.^8^ Furthermore, the efficacy of TNF inhibitors has been associated with genetic variants at the 6q23.3 susceptibility locus, where *TNFAIP3* encodes A20, an inhibitor of TNF signalling.^43^ The current study establishes two new psoriasis susceptibility loci harbouring genes encoding therapeutic targets (Supplementary Table 16). We found novel psoriasis susceptibility variants centred on the 5’ untranslated region of *IL17RA* at chromosome 22q11.1 (lead variant rs917864, OR: 1.08, P=3.9×10^−9^). The interleukin-17 receptor A subunit encoded by this gene^44^ is bound with high affinity by brodalumab, a biologic therapy demonstrated to confer effective long-term control of psoriasis.^45, 46^ Similarly, the aryl hydrocarbon receptor is encoded by *AHR*, the closest protein-coding gene to the novel 7p21.1 association signal (lead variant rs78233367, OR: 1.06, P=1.3×10^−8^). Activation of this receptor by the recently approved topical AHR- modulating agent Tapinarof effectively controls psoriasis symptoms.^47–49^

In addition to AHR, a series of other transcription factors were implicated as putative causal genes by our functional fine mapping approach. We observed examples of newly reported association signals mapping to fundamental processes of cell differentiation, proliferation and trafficking that contribute to established psoriasis pathomechanisms including interferon response, T cell regulation and keratinocyte hyperproliferation. The elongation factor gene ELL, which maps to the newly identified 19p13.11 locus, is predicted by TWAS to be upregulated in blood in the presence of psoriasis-associated alleles. Moreover, by stabilizing RNA polymerase II within the super elongation complex, ELL has been demonstrated to help sustain the expression of the epidermal proliferation genes that are greatly upregulated in psoriasis.^50^ A deleterious missense variant in *POU2F3* (rs7110845, p.His154Arg, P=2.3×10^−13^), which encodes an epidermal transcription factor critical to keratinocyte differentiation,^51^ is reported here for the first time. CCAAT enhancer binding transcription factors, implicated in myeloid lineage commitment and regulation of inflammatory cytokines,^52–54^ are encoded by *CEBPB* at the known 20q13.13 locus, and *CEBPA* and *CEBPG* at the newly reported 19q13.11 locus. The latter gene is predicted by TWAS to be downregulated in sun-exposed and unexposed skin in the presence of psoriasis risk variants and would therefore be consistent with previous reports that C/EBPγ suppresses proinflammatory cytokine activity.^55^

Transcriptional regulation of interferon responses is further coordinated by genes at a series of psoriasis susceptibility loci. The interferon regulatory factor genes *IRF1* and *IRF5* are predicted by TWAS to be upregulated in sun-exposed skin and across tissues, respectively, in the presence of psoriasis risk variants at the known 5q31.1 (*IRF1*) and newly reported 7q32.1 (*IRF5*) loci. We also identified a susceptibility locus at 20q13.2 that contains *NFATC2*, encoding a transcription factor that interacts with interferon regulatory factor 4 (encoded by *IRF4* at the 6p25.3 locus) to regulate T cell development via enhanced IL-4 expression. Epigenetic regulation of interferon signalling is implicated through the histone acetyltransferase genes *KAT5* (11q13.1 locus) and *KAT8* (16p11.2), which regulate either directly or indirectly the transcriptional activity of IRF3^56, 57^, and are predicted to be upregulated in skin.

We shed further light on the cellular context of the interferon response in psoriasis through our single-cell expression data. Consistent with its role in in epidermal proliferation^50^, we found that the *ELL* gene described above is a TWAS candidate predominantly expressed in keratinocytes (Figure 3). Another cluster of TWAS target genes, including many innate immune genes (e.g., *DHX58, STAT2, TYK2*) are highly expressed in melanocytes from psoriasis lesions. In addition to producing melanin for photoprotection, melanocytes assume an activated phenotype in psoriasis lesions^58^ and express functional toll-like receptors TLR2, 3, 4, 7 and 9^59^, enabling innate antibacterial and antiviral immune responses. Melanocytes have also been recently proposed as target cells of the HLA-C*06:02-restricted autoimmune response in psoriasis via the melanocyte autoantigen ADAMTSL5, which could provide a skin-specific target for autoimmune attack.^60, 61^ More generally, our single-cell expression analysis suggests that psoriasis genetic risk is mediated by a range of cell types beyond the established triad of T cells, dendritic cells and keratinocytes.

We found predicted deleterious protective variants in two genes encoding members of the RIG-I like receptor (RLR) family, linking recognition of viral RNAs to interferon signalling^62, 63^: *DHX58* at chromosome 17q21.2 and *RIGI* (formerly *DDX58*) at 9p21.1 (Supplementary Table 7). Prior to this study, *STAT3* had been considered the leading functional candidate in 17q21.2 due to its prominent role in IL-23 signal transduction.^10^ Previous reports have implicated deleterious coding variants in *IFIH1*, the third member of the RLR family, as conferring protection against psoriasis^8, 64^ and psoriatic arthritis^65^; the protective *IFIH1* variants rs35667974 and rs1990760 are both highly significant in the current meta-analysis (P=5.0×10^−23^ and 7.0×10^−50^, respectively) but not prioritised in credible sets due to insufficient effective sample size and the presence of a more strongly associated intronic variant (rs2111485), respectively. While the *RIGI* and *DHX58* coding variants both show a low posterior probability of being causal (the intronic variant rs11795343 being strongly preferred in *RIGI*, with posterior probability 0.922), the fact that potentially deleterious coding variants are found to be associated with reduced psoriasis risk in all three known human RLR family members is intriguing. These findings are consistent with reports of rare gain-of-function mutations in *RIGI* and *IFIH1* underlying Singleton-Merton syndrome, in which psoriasis is a clinical feature.^66, 67^ Much remains to be learned about how the role of RLR family members in driving interferon signalling is affected by psoriasis-associated genetic variation.

This study represents a major advance in our understanding of the genetic basis of psoriasis. The number of documented psoriasis susceptibility signals has approximately doubled, with better refinement of known loci to highlight plausible biological mechanisms through which they influence psoriasis risk. We propose novel disease mechanisms, including a role for the disruption of basic cellular machinery such as transcription and epigenetic modulation in regulating the inflammatory process in psoriasis. For the first time, genetic susceptibility signals are contextualised to specific skin cell types and cytokine signalling pathways. Our data point to the participation in psoriasis pathogenesis of previously underappreciated cell types such as melanocytes. This work will underpin the next era of molecular studies in psoriasis and psoriasis therapeutics.

## Methods

### Contributing GWAS studies

Genotype data for chronic plaque psoriasis cases and unaffected or population-based controls were compiled for 18 contributing studies, each of which underwent stringent QC at one of eight contributing analysis centres (Supplementary Table 17). Full details are provided in Supplementary Methods. Prior to association testing, inter-dataset duplicated and first- or second-degree related participants were identified using KING (version 2.0)^68^ by sharing subsets of between 2,502 and 6,864 genotyped markers outside known psoriasis-associated regions. After removing duplicated and related samples, the final sample size was 36,466 cases and 458,078 controls. The cumulative effective sample size was 103,614 (Supplementary Table 17). All participants were of European ancestry. The contributing analysis centres were responsible for genome-wide imputation and association testing (further details in Supplementary Methods).

### Meta-analysis

Summary statistics from individual studies were aligned based on GRCh37 positions and alleles; reference alleles were checked for consistency across studies. In many datasets, more than one imputation reference panel was used to maximise imputation quality;^9^ summary statistics for each variant were preferentially taken from the version with highest imputation quality (INFO or R^2^) score. Standard error-weighted fixed effects meta-analysis was performed using METAL v2020-05-05.^69^

Genomic inflation was calculated based on a set of 170,786 LD-independent variants outside of previously established psoriasis susceptibility loci having minor allele frequency > 0.05 (Supplementary Methods).^20^ LD score regression intercept was calculated using LDSC software with default settings and precomputed LD scores derived from 1000 Genomes data.^21^

### Associated regions and independent signal identification

Associated genomic regions were identified using Genome-wide Complex Trait Analysis conditional and joint analysis (GCTA-COJO).^26^ To facilitate this, the autosomes were first partitioned into distinct LD blocks (Supplementary Methods). A region (LD block) was considered associated with psoriasis susceptibility if at least one variant tested in three or more contributing studies and with N_eff_ > 10,000 (11,808,957 eligible variants in total) achieved genome-wide significance (P<5×10^−8^). The LD block that includes the MHC region (chr6, 24.0-36.3Mb) extends beyond the established boundaries of the extended MHC (chr6, 25.7-33.4Mb^70^); we observe an association outside of the established boundaries that could plausibly be driven by weak LD with the strongly associated MHC variants (Supplementary Figure 1).

For the identification of additional independent association signals within associated regions we used a more stringent subset of variants having N_eff_ > 93,252 (90% of maximum possible), with five non-MHC regions omitted due to lack of variants meeting this sample size threshold. We employed GCTA-COJO with a custom reference panel to determine independently associated lead variants using a stepwise model selection procedure, and to estimate conditional association statistics for each signal in LD blocks with multiple independent signals (Supplementary Methods). We were unable to estimate additional independent signals within the MHC block (Supplementary Methods).

To annotate our genomic regions against psoriasis susceptibility loci previously established at genome-wide significance in European populations or other ancestries, we reviewed recent GWAS meta-analyses^7–9, 22–25, 71^ and assessed all psoriasis associations in GWAS Catalog (accessed 31 October 2022).^72^

We used LDSC^21^ to estimate the total common SNP heritability for psoriasis on the liability scale, assuming a population prevalence of 1.5%. We note that the heritability estimated by LDSC when including the MHC region is to be interpreted with caution due to its complex genetic architecture.^21, 73^ Variance explained by our genome-wide significant associations was estimated using the Mangrove package in R,^74^ based on the jointly estimated effect sizes from COJO and with allele frequencies estimated from 1000 Genomes data. Since independent signals were not estimated in the LD block containing the MHC region, only the marginal effect size of the lead SNP was included, which is likely to underestimate the heritability attributable to this region.

### Statistical fine-mapping

For each independent association signal, we used the method suggested by Wen and Stephens,^75^ and implemented in previous work,^22^ to calculate posterior probabilities (PP) for each variant being causal. Within each LD block, PPs were calculated for variants with N_eff_ > 90% of maximum, using the joint meta-analysis association p-values estimated by GCTA-COJO to account for the presence of multiple association signals. Bayesian 95% credible sets were subsequently constructed by incorporating variants in decreasing PP order until the cumulative PP exceeded 0.95. Since independent signals could not be established for the LD block containing the MHC region, the 95% credible set for the lead signal was based on unconditional association statistics. We did not estimate credible sets for five signals where no variants had N_eff_ > 90% of maximum.

We also assessed improvement in statistical fine-mapping relative to the GWAS datasets available in the previous (2017) psoriasis GWAS meta-analysis.^7^ To this end, we selected 63 psoriasis susceptibility loci that were genome-wide statistically significant in either the 2017 meta-analysis and/or other published studies of white European ancestry populations and, for comparability, re-computed associations for these loci in the 2017 meta-analysis without the PAGE dataset (which was typed using the Immunochip and therefore lacks full genome-wide coverage). For the comparison, 95% credible sets were calculated in both the current and (re-computed) 2017 meta-analysis within windows of 200kb around the 2017 lead markers, based on unconditional association statistics; credible set construction was restricted to variants well-imputed for a N_eff_ > 90% of the maximum possible meta-analysis sample size.

### Prioritisation of causal variants

For variants identified in 95% credible sets (5,344 distinct variants), we assessed the potential contribution to disease risk via altered protein-coding or regulation. Variants with predicted protein-altering consequence were identified using the Variant Effect Predictor,^76^ along with associated Combined Annotation Dependent Depletion (CADD) scores.^77^ To identify regulatory variants likely to influence psoriasis risk we calculated generic and tissue-specific regulatory prediction scores with the RegulomeDB-based method TURF.^27^ The TURF analyses used default settings and reference data, and excluded indels (5,102 SNPs in total). For each independent association signal, we checked for prioritised regulatory variants, which we defined as variants with: (i) generic regulatory probability >0.7, (ii) at least 50% share of generic regulatory probability among variants within their credible set, and (iii) no other credible set variant having generic regulatory probability >0.5. We assessed blood- and skin-specific regulatory probabilities for prioritised variants and compared these to 49 other tissues with tissue-specific regulatory probabilities.

### Transcriptome-wide association study

Predictions of differential gene expression in the presence of psoriasis-associated genetic variation were generated using the S-PrediXcan method^78^ implemented in the Complex Trait Genetics Virtual Lab (CTG-VL).^79^ We predicted expression differences in whole blood (n = 6,275 genes with prediction models available), sun-exposed skin (n = 9,085) and sun-unexposed skin (n = 7,617) using GTEx v7 reference data.^80^ We employed a Bonferroni-adjusted transcriptome-wide significance threshold of 2.18×10^-6^ (22,977 tests performed) to assess differential expression associated with psoriasis.

To establish the physical location of psoriasis-associated TWAS genes relative to susceptibility signals, we considered all protein-coding and long non-coding RNA genes from Ensembl (GRCh37 version 109).^81^ Genes were allocated to psoriasis-associated genomic regions based on having transcription start site (TSS) within the corresponding LD block, or within 1Mb of the lead variant. Gene distances in each region were compared based on the distance from TSS to the lead psoriasis-associated variant. Significant TWAS genes outside of psoriasis-associated genomic regions were mapped based on distance to the variant within 1Mb having lowest meta-analysis p-value (all psoriasis-suggestive variants, P_meta_<10^-5^).

### Single-cell functional analyses

To delineate the specific skin cell types underlying psoriasis-associated TWAS genes, we studied their expression profiles using scRNA-seq data that we recently reported.^82^ For each TWAS gene, we computed the average expression in each of 12 cell types derived from non-lesional or lesional skin of psoriasis cases (24 biopsy type/cell type combinations in total; n_non-lesional_=11, n_lesional_=14), and subsequently calculated the standardized expression across cell types for each gene. We grouped genes with consistent expression profiles and identified five clusters using hierarchical clustering.

We also investigated the overlap between psoriasis-associated TWAS genes and transcriptomic changes induced by psoriasis-linked cytokines. Specifically, gene expression profiles were generated for keratinocytes from 50 healthy adult donors before and after stimulation with a range of cytokines (IL-4, IL-13, IFN-α, IFN-γ, TNF-α, and IL-17A), as described previously.^36, 83^ We compared and tested for statistical enrichment (hypergeometric test) our list of non-MHC TWAS genes against the top 90 most statistically significant upregulated genes in each keratinocyte cytokine challenge.

### Biological pathway analysis

We used DEPICT v1.1 to identify sets of genes with coordinated function in which genes from psoriasis susceptibility loci are highly expressed.^34^ As input we used a list of 214 independent and genome-wide significant variants derived from the full meta-analysis summary statistics via distance- and LD-based clumping (r^2^>0.05 within 2Mb windows) in PLINK^84^ based on 1000 Genomes European samples,^85^ except for the extended HLA region where only the lead SNP was included. For comparison purposes, we repeated the analysis using a subset of 158 variants within 1Mb of previously established psoriasis susceptibility loci. In each case, functional gene sets were considered significantly enriched at a false discovery rate (FDR) of <0.05 based on 14,462 gene sets tested.

### Correlation and causation analyses

Genetic correlations^86^ were estimated in CTG-VL using summary statistics for an initial set of 1,376 traits (CTG-VL default list), a majority of which were derived from UK Biobank by the Neale lab (http://www.nealelab.is/uk-biobank). We excluded traits that were too general or non-specific, represented follow-up questions for specific groups (e.g., details of presentation for severe mental illnesses) or detailed lifestyle questions (e.g., specifics of diet or occupation), psoriasis and duplicated or otherwise difficult-to-interpret traits, resulting in 592 traits categorised into disease and health (n=345), physical and functional measures (n=210) or lifestyle and quality of life (n=37) (Supplementary Table 14). Significant genetic correlation was identified among these traits based on a Bonferroni-adjusted p-value threshold of 8.45×10^−5^ (592 tests performed), and we checked that all significant traits had heritability z-score >4.

Partial genetic causality between psoriasis and a range of other traits was estimated using a latent causal variable (LCV) model^41^ implemented in CTG-VL. Of the 592 traits for which genetic correlation was assessed (above), LCV results were available for 585. We focused on 108 traits with significant genetic correlation at FDR<0.05. Traits with evidence for a causal relationship were subsequently identified based on a significant genetic causality proportion (FDR<0.05 among 108 traits).

## Data Availability

Data in the present study will be made publicly available on publication of the manuscript in a peer-reviewed journal.

## Acknowledgements

The BSTOP steering committee comprises: Professor David Burden (Chair), Professor Catherine Smith, Professor Jonathan Barker, Professor Sara Brown, Dr Nick Dand, Dr Satveer Mahil, Helen McAteer, Dr Julia Schofield and Professor Stefan Siebert. The Estonian Biobank research team comprises: Tõnu Esko, Andres Metspalu, Lili Milani, Reedik Mägi and Mari Nelis.

This research has been conducted using the UK Biobank Resource (approved project 15147). It uses data provided by patients and collected by the NHS as part of their care and support. Copyright © 2023, NHS England. Re-used with the permission of NHS England and UK Biobank. All rights reserved.

We gratefully acknowledge the contributions of patients and family members, other research participants and clinical staff involved in recruitment.

## Author Contributions

J.T.E. and M.A.S. jointly supervised the study, which was conceived by J.T.E. and J.N.B. N.D. L.C.T., M.A.S. and J.T.E. contributed to study design. N.D., A.B., V.C., T.E., J.F., A.F., D.D.G., W.G., U.H., K.K., S.K., W.L., M.L., R. Mägi, R.P.N., P.R., A. Reis, C.H.S., J.N.B. and J.T.E. coordinated individual GWAS studies, with data preparation and association testing performed by N.D., P.E.S., J.B., D.E., J.N., M.T-L., L.F.T., T. Traks, S.U. and L.C.T. The meta-analysis was performed by N.D., with downstream bioinformatic analysis by N.D., P.E.S., J.R.S. and L.C.T. J.E.G. and L.C.T. generated transcriptome data. P.D.M. and J.T.E. led the biological interpretation of results. G.A., D.B., F.B., A.C.B., M.A.B., H.B., F.C., R.C., C.J.C., M.D., E.E., O.F., S. Gerdes, C.E.M.G., S. Gulliver, P. Helliwell, P. Ho, P. Hoffmann, O.H., Z.H., K.H., D.J., M.K., C.K., C.L., S.H.L., F.M., S.K.M., N.M., R. McManus, E.H.M., M.J.N., M.N., V.O., J.R.O., M.T.P., B.E.P., A. Ramming, J.R., C.R., M.K.S., G.S., B.S., T. Tejasvi, H.T., J.J.V., E.M.W., R.B.W., R.W., S.W., X.W. and Z.Z. contributed to the acquisition, generation and/or processing of data for individual GWAS or transcriptome studies. N.D., P.E.S., P.D.M., M.A.S. and J.T.E. drafted the manuscript and all authors critically reviewed the manuscript.

