## Supplementary Material for "GWAS meta-analysis of psoriasis identifies new susceptibility alleles impacting disease mechanisms and therapeutic targets"

**Supplementary material
for**

**GWAS meta-analysis of psoriasis identifies new susceptibility alleles impacting disease mechanisms and therapeutic targets**

**Supplementary Methods**

*Contributing GWAS studies*

The 18 contributing studies included 15 datasets with psoriasis cases ascertained via dermatologists or other secondary care specialist physicians (five previously analysed^1^: CASP, ExomeChip, Genizon, PsA and WTCCC2; ten newly genotyped: BSTOP, Erlangen, Estonia, Kiel, Manchester, Michigan, Newfoundland, Scotland, Toronto and UCSF) and three datasets derived from biobanks (Estonian Biobank, HUNT and UK Biobank). Details of ethics approval, ascertainment, genotyping, quality control, genome-wide imputation, and association testing for the 18 contributing studies are provided separately in the Supplementary Methods Table.

*LD reference panel for statistical fine-mapping*

For GCTA-COJO signal identification and fine-mapping we employed a custom LD reference panel comprising genome-wide well-imputed (Mach r^2^ ≥ 0.7) data from six GWAS datasets: CASP, ExomeChip (i.e., including full GWAS content), Genizon, PsA and WTCCC2 from this study, plus previously analysed data from Kiel^2^ that was omitted from the current meta-analysis because most samples had been re-genotyped as part of the larger Kiel dataset included here. The reference panel comprised a total of 24,069 samples, and included all variants with an effective sample size N_eff_ > 90% of maximum for the full meta-analysis.

This reference panel was also used to identify a set of 170,786 independent markers for estimation of genomic inflation. These were identified using pairwise LD-pruning in PLINK,^3^ using window size 1,500 Kb, step size 150 variants and R^2^ threshold 0.2.

*Partitioning of genome into distinct LD blocks*

We used an optimal procedure^4^ to partition the autosomal genome into 1,703 approximately independent LD blocks. The LD reference was based on a curated version of 1000 Genomes Project genotypes^5-7^ from which we extracted 503 unrelated samples of European ancestry and excluded variants with minor allele frequency < 0.05. The minimum and maximum allowable number of LD reference variants in each LD block was set to 200 and 10,000, respectively. For each autosome, the maximum number of blocks to consider was chosen to match publicly available results of an older, widely-adopted genome-partitioning method applied to European-ancestry samples.^8, 9^ Compared to application of the European-based block boundaries from this older method to our LD reference, we found that block boundaries generated by the optimal method yielded a 600-fold reduction in the cost of splitting autosomes, where cost is defined as the sum of squared correlations between variants from different blocks, restricting to all r^2^ > 0.05.

*Identification of secondary association signals*

For the identification of additional independent association signals within associated regions we used a stringent subset of variants having N_eff_ > 93,252 (90% of maximum possible) and employed GCTA-COJO, version 1.93.3-beta.^10^ First, we determined independently associated lead variants using a stepwise model selection procedure (--cojo-slct procedure). For LD blocks with multiple independent signals, we further estimated association statistics for each independent signal conditioned on the other independent signals in the same LD block (-‑cojo-cond procedure). All analyses used default parameter values (--cojo-p 5e-8, --cojo-wind 10000, cojo-collinear 0.9, --diff-freq 0.2). We do not report results of this procedure for the MHC block, for two reasons: (i) testing in a subset of datasets for which genotype data were available showed very poor consistency between COJO pseudo-conditional analysis and “gold-standard” genotype-based conditional analysis, likely due to the complex LD structure in the region, suggesting that the COJO-based model selection procedure does not reliably identify independent MHC signals; (ii) a substantial fraction of variants within the MHC region failed to achieve the minimum imputation quality threshold in the HUNT study, leading to few variants having N_eff_ > 93,252 (90% of maximum possible).

**Supplementary Note**

*Definition and number of psoriasis susceptibility loci*

We defined a psoriasis susceptibility locus as an LD block (see Supplementary Methods) in which at least one variant exhibited genome-wide significant evidence of association with psoriasis (P<5×10^‑8^) in the full meta-analysis. We considered a psoriasis susceptibility locus (LD block) to have been previously reported in Europeans if it contained the lead variant for one of the 65 association signals reported by previous studies.^1, 11, 12^ Note that two of the previously reported association signals did not reach genome-wide significance in the current meta-analysis (as highlighted in main results) and that four of our loci (LD blocks) contained lead variants for two previously reported association signals (Supplementary Table 2). Therefore, 63 of the 65 previously reported association signals map to 59 loci in the current study. We found a further 50 susceptibility loci (LD blocks) that did not overlap with any of the 65 previously reported association signals. We consider these loci to be newly reported in European-ancestry populations. The 59 previously reported and 50 newly reported loci give a total of 109 susceptibility loci in Europeans. One of these loci encompasses the MHC region, which has been shown to include multiple independent association signals^13^ and which we did not interrogate further in this study. We found that 27 of the remaining 108 loci contained multiple independent susceptibility signals, including all four loci that mapped to two of the 65 previously reported association signals.

stylefix

**Supplementary Figure 1 – Association results across the extended MHC region**

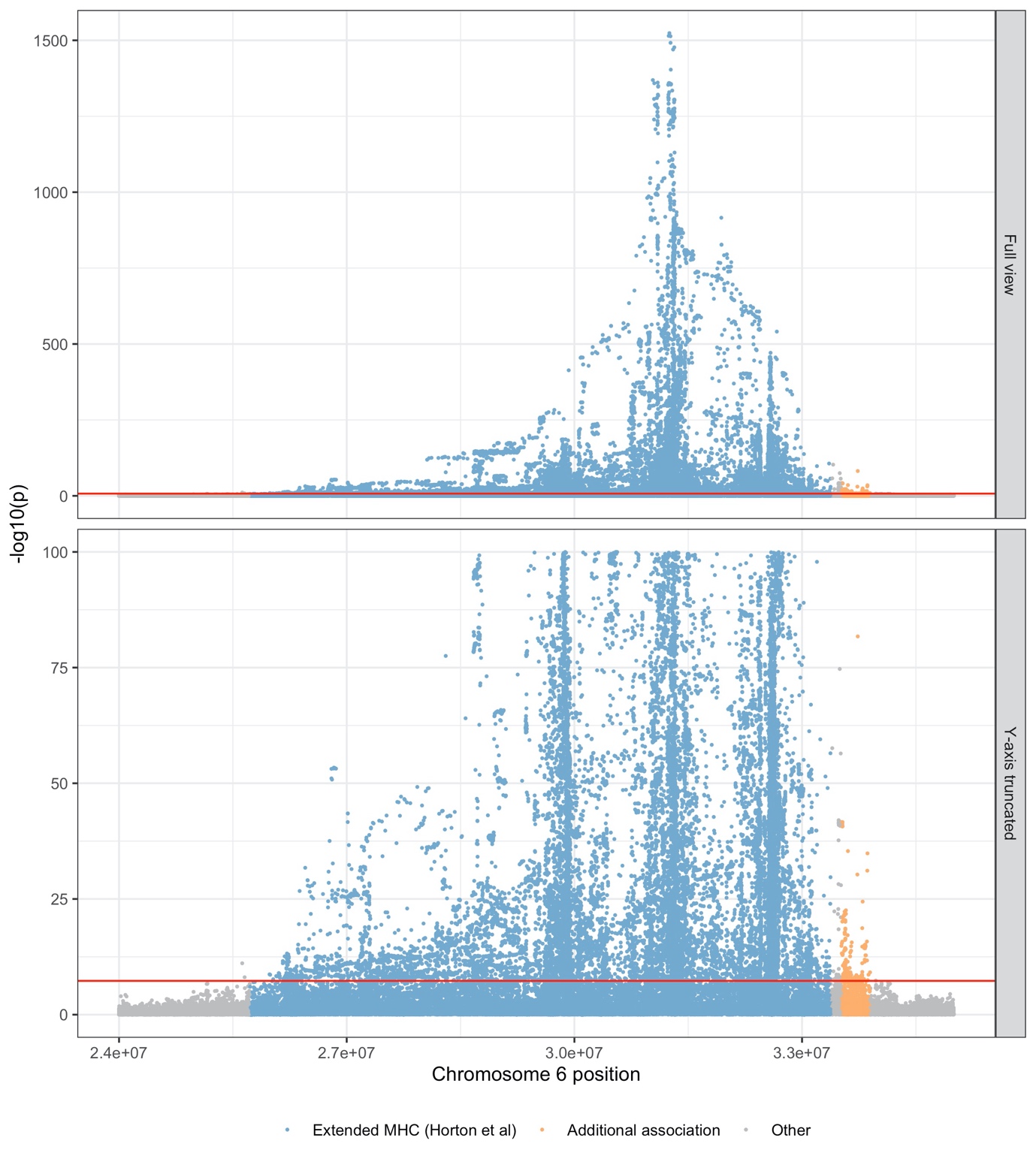

x-axis, position on chromosome 6; y-axis, -log10(P-value) of association; blue points, variants within the extended MHC region as defined by Horton et al. (Nat Rev Genet, 2004); orange points, variants in an adjacent region of genome-wide significant association outside the Horton-delimited MHC. These signals all occur within the same LD block and as such are counted as a single susceptibility locus, although the MHC is known to harbour multiple independent psoriasis susceptibility signals.

**Supplementary Figure 2 – Comparison of 95% Bayesian credible set length to previous GWAS meta-analysis**

Comparison of 95% Bayesian credible set lengths to previous GWAS meta-analysis. Each point represents a different association signal established in the previous meta-analysis (Tsoi et al., 2017). Point colour indicates direction of change, blue dashed line indicates equality.

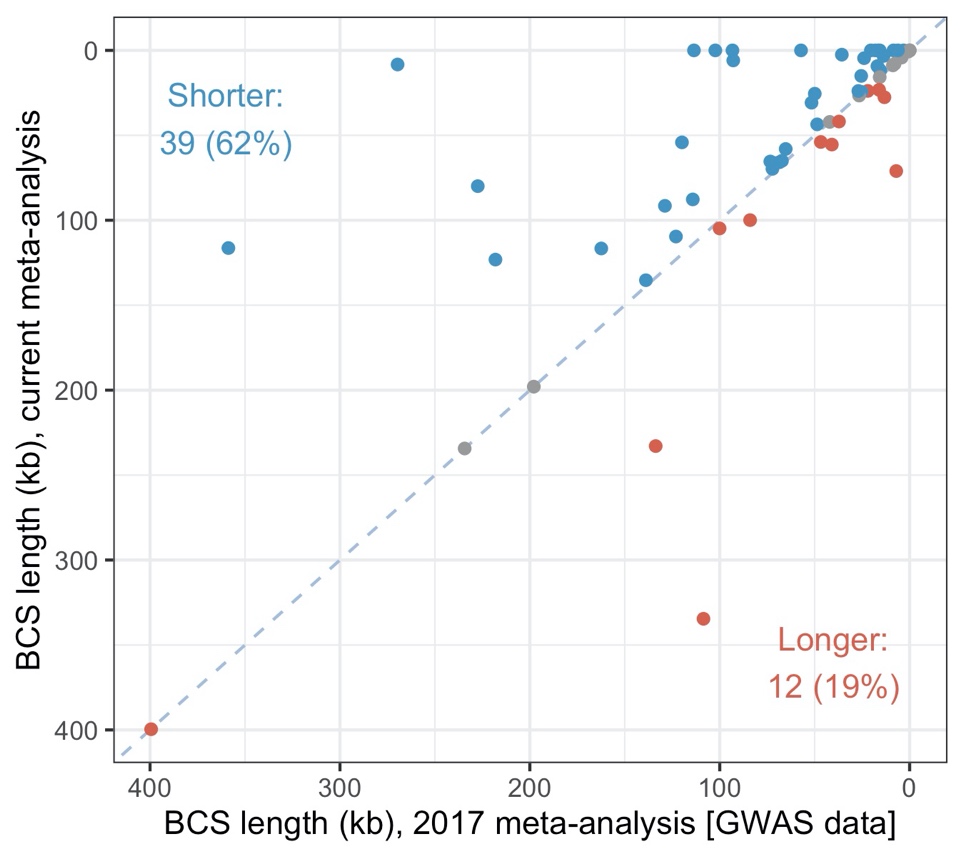

**Supplementary Figure 3 – Credible set enrichment for predicted regulatory variants**

Variant regulatory probabilities estimated by TURF (y-axis), summarised by size of Bayesian credible set (x-axis). Upper boxplot: TURF generic probabilities; lower boxplot: maximum of skin-specific and blood-specific regulatory probability is used for each variant. Boxes represent interquartile range (IQR) with horizontal bar at median. Whiskers indicate full range of data except for outlying variants (>1.5× IQR from box) marked individually. BCS, Bayesian credible set.

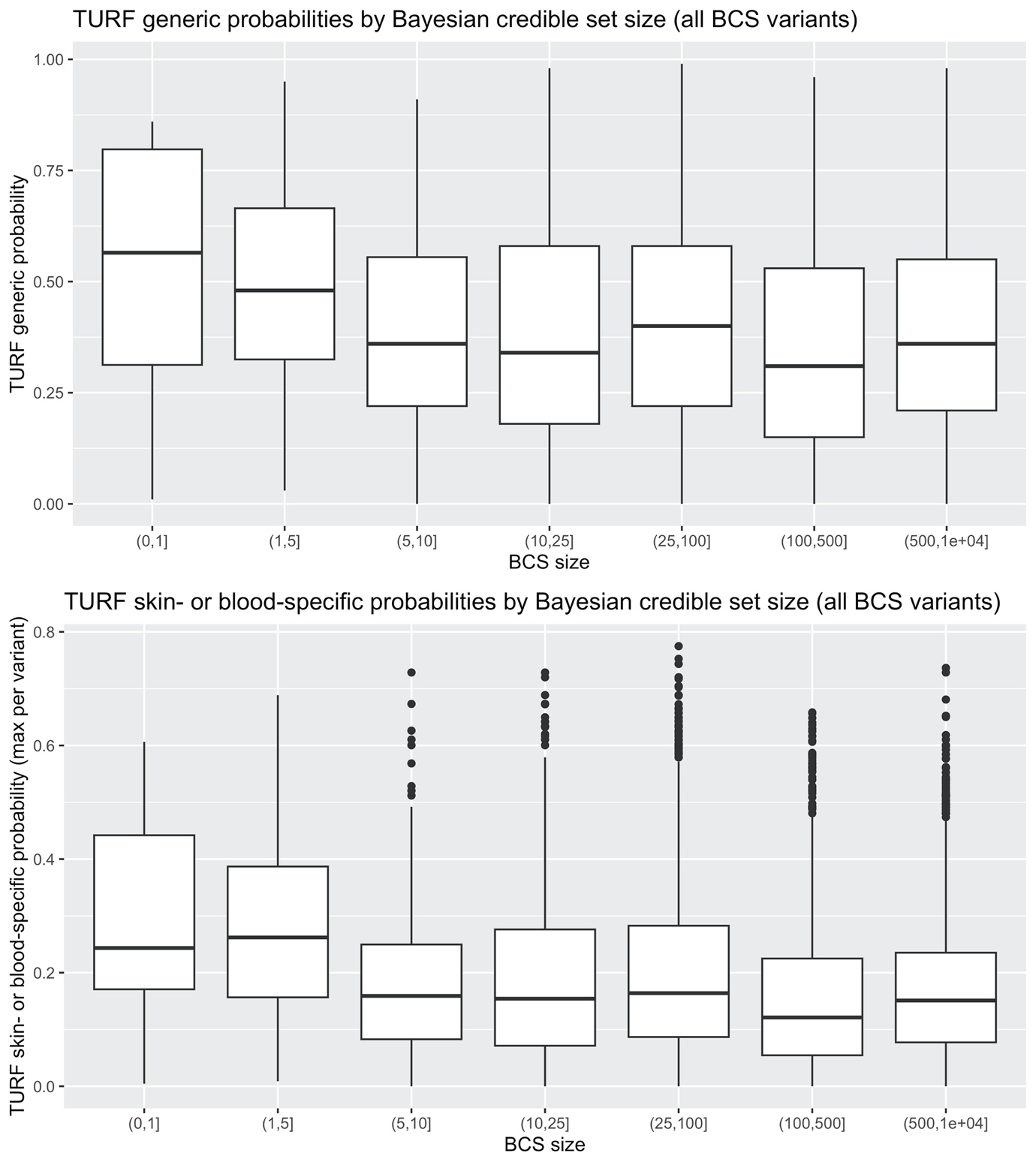

**Supplementary Figure 4 – High-confidence regulatory variants identified by TURF analysis**

For the 14 candidate regulatory variants, plots show generic (black points) and tissue-specific (grey) regulatory probabilities (y-axis) estimated by TURF for all tissues (x-axis). Blood and skin are highlighted in orange and blue, respectively (inverted triangles for ease of identification).

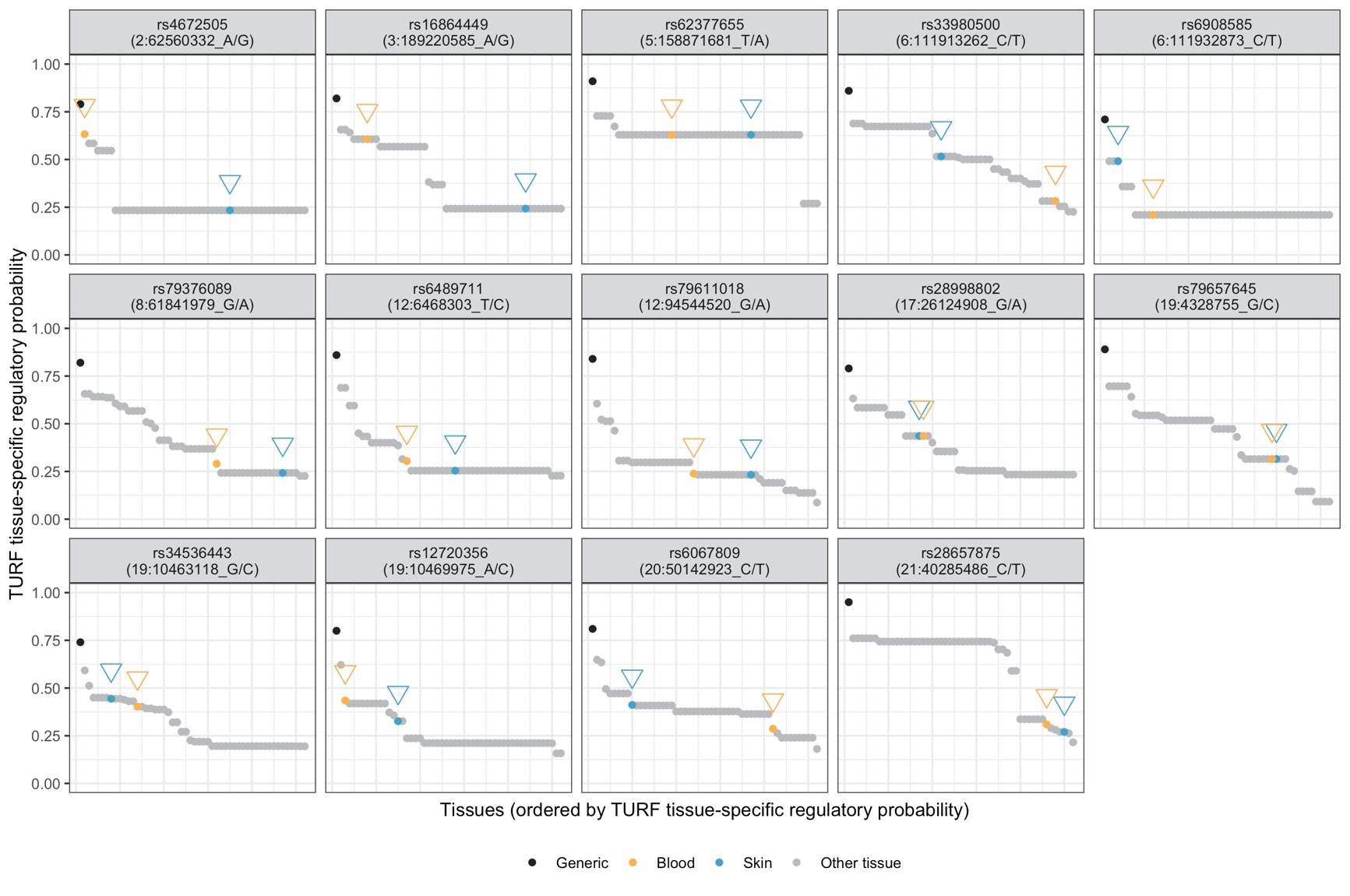

**Supplementary Figure 5 – Manhattan plots for transcriptome-wide association study**

Each point represents a gene with predicted expression; point colours alternate by chromosome with MHC region genes on chromosome 6 greyed out; x-axis: position in genome; y-axis: -log_10_(TWAS association p-value); horizontal orange line: transcriptome-wide significance threshold (3.9×10^-6^); panels represent predicted expression results in different tissues (sun-unexposed skin, sun-exposed skin, whole blood).

**A** – All results

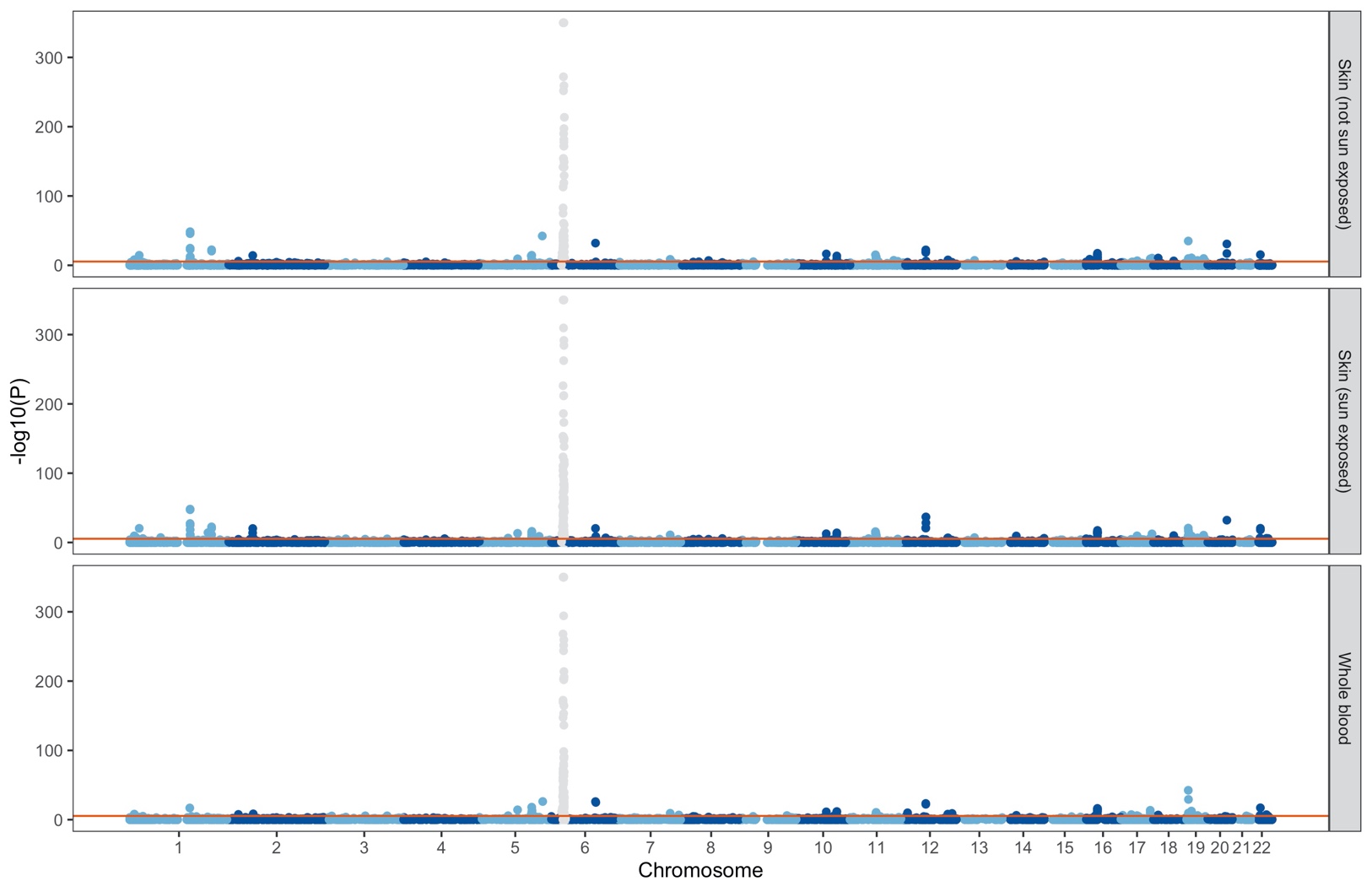

**B** – Truncated y-axis for clearer resolution at non-MHC signals

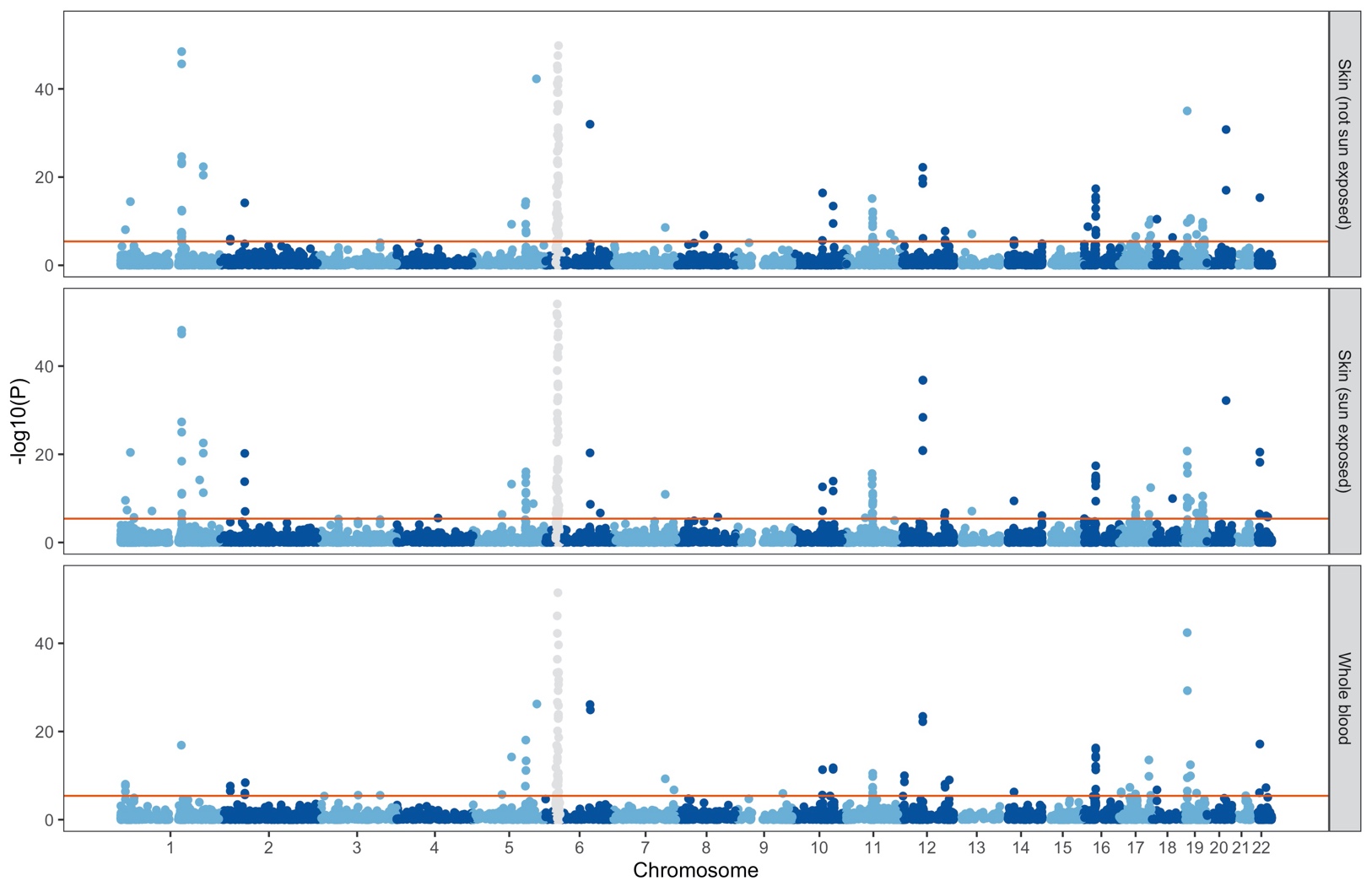

**Supplementary Figure 6 – Physical location of significant TWAS genes**

Genes allocated to each of 108 non-MHC psoriasis susceptibility regions (see Methods) are plotted by position relative to the lead meta-analysis variant; x-axis: position relative to lead variant; y-axis: one row per genomic region; vertical tick marks indicate lower and upper limits of each LD block; orange points indicate transcriptome-wide significant genes; biotype “other” refers to a handful of TWAS genes that are transcribed or processed pseudogenes or encode lincRNAs.

**A** – LD blocks 1-36

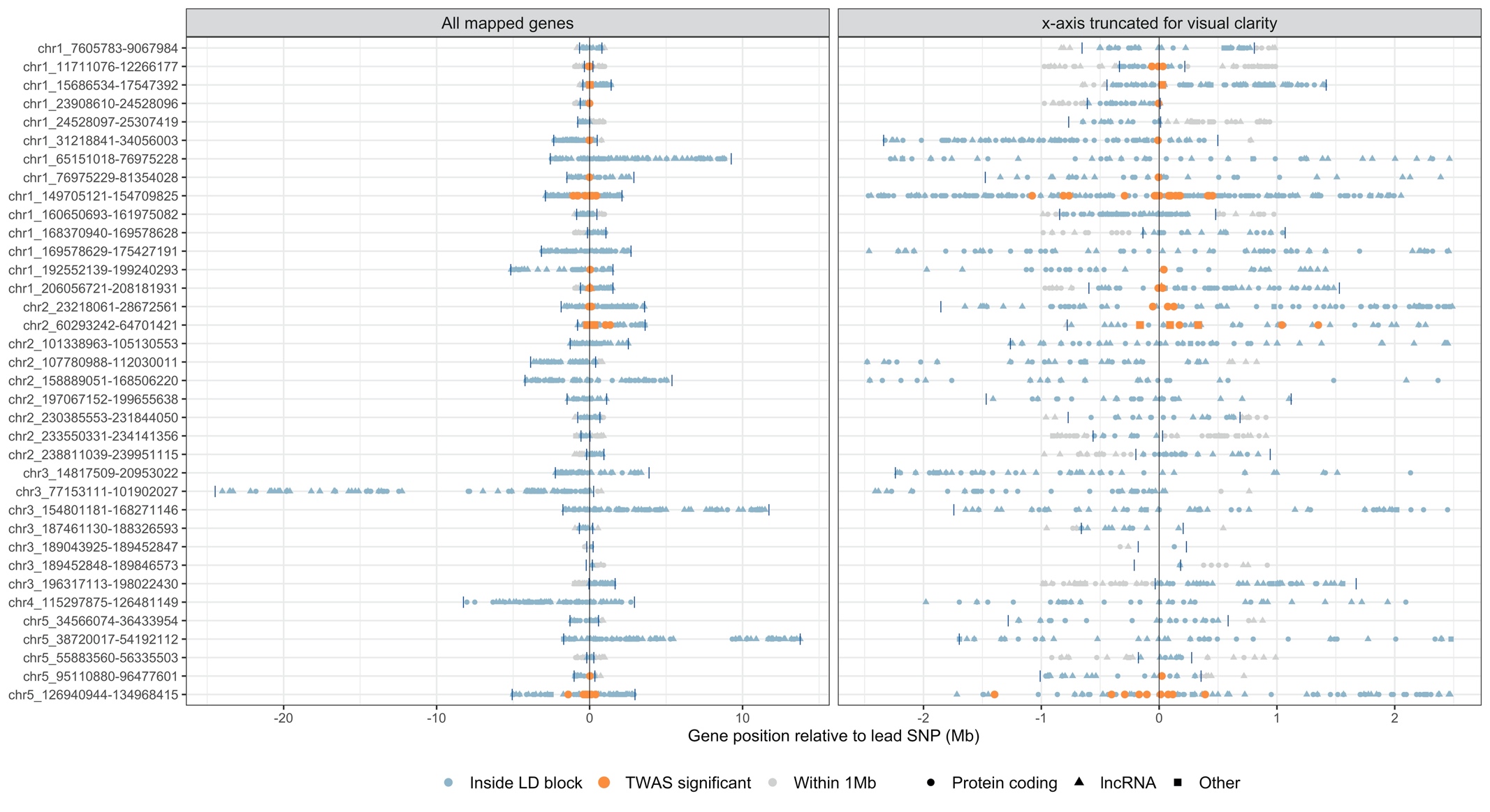

**B** – LD blocks 37-73 (excludes MHC block 41)

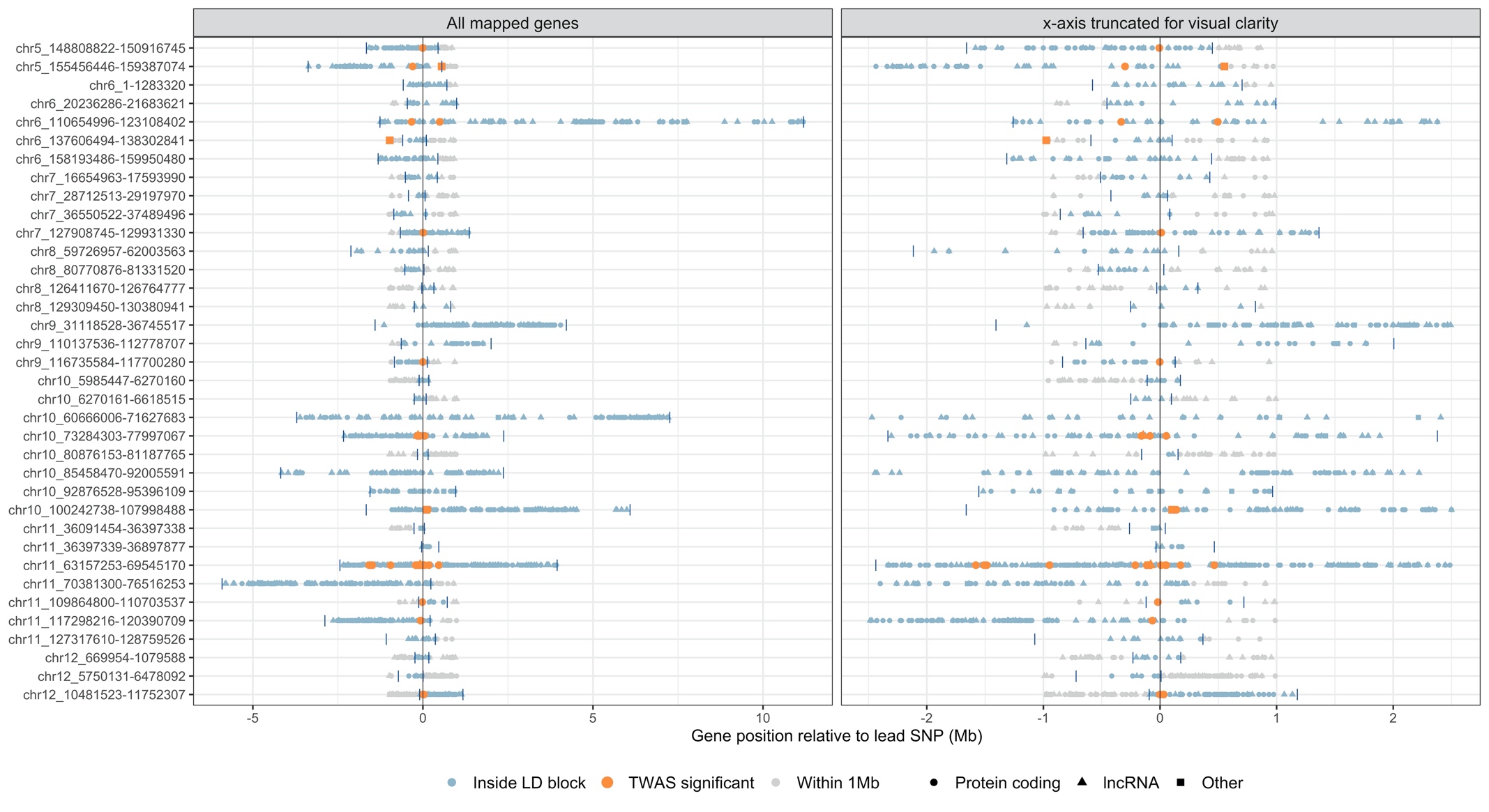

**C** – LD blocks 74-109

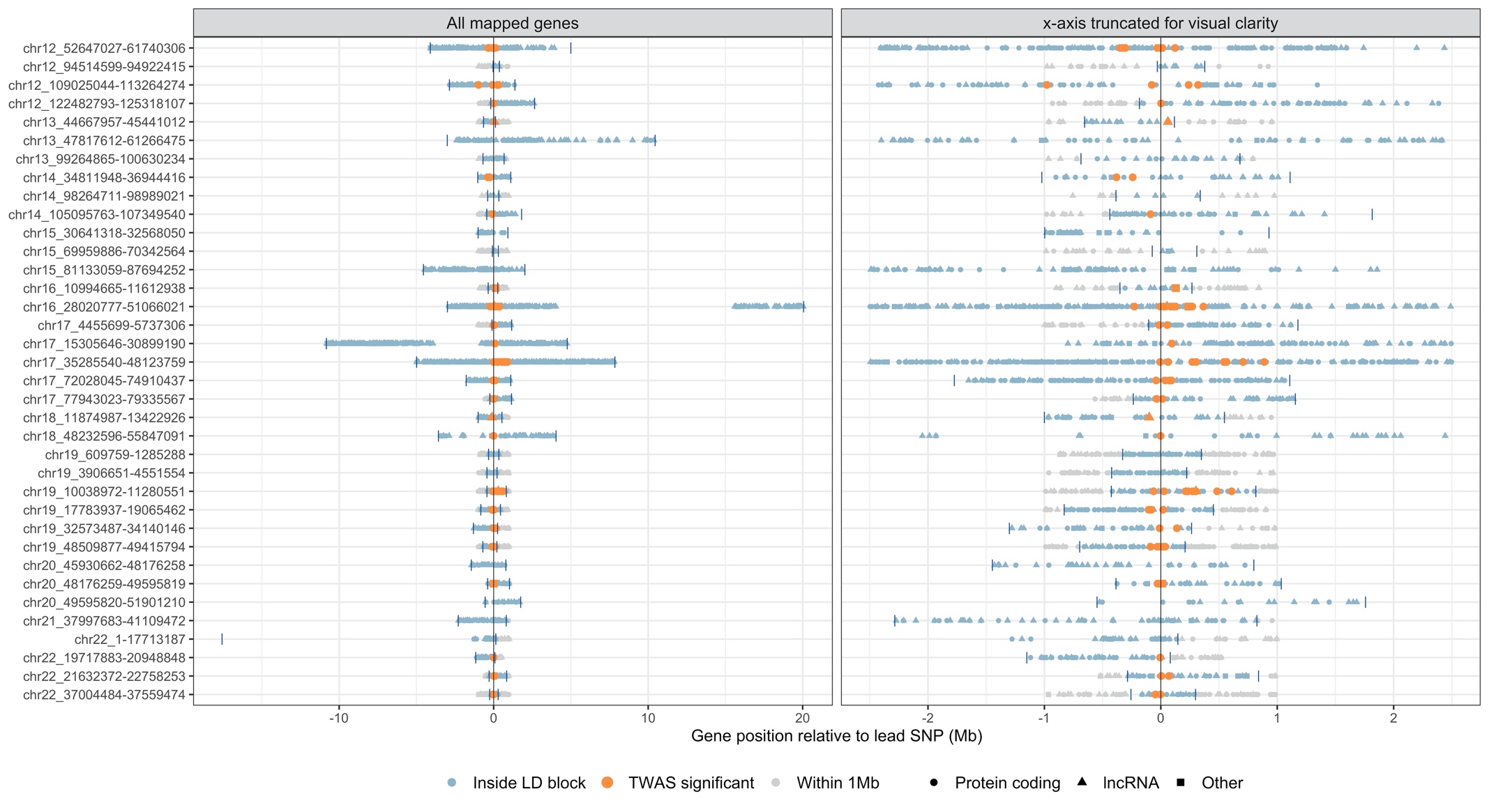

**Supplementary Figure 7 – Comparison of gene set enrichment results from DEPICT**

Each point represents a gene set; x-axis, rank among all gene sets (A, left-hand plot) or ‑log_10_(enrichment p-value) (B, right-hand plot) based on DEPICT analysis using previously reported psoriasis susceptibility regions only; y-axis, rank (left-hand plot) or ‑log_10_(enrichment p-value) (right-hand plot) based on DEPICT analysis using all psoriasis susceptibility regions; point colour indicates enrichment status based on false-discovery rate <5%.

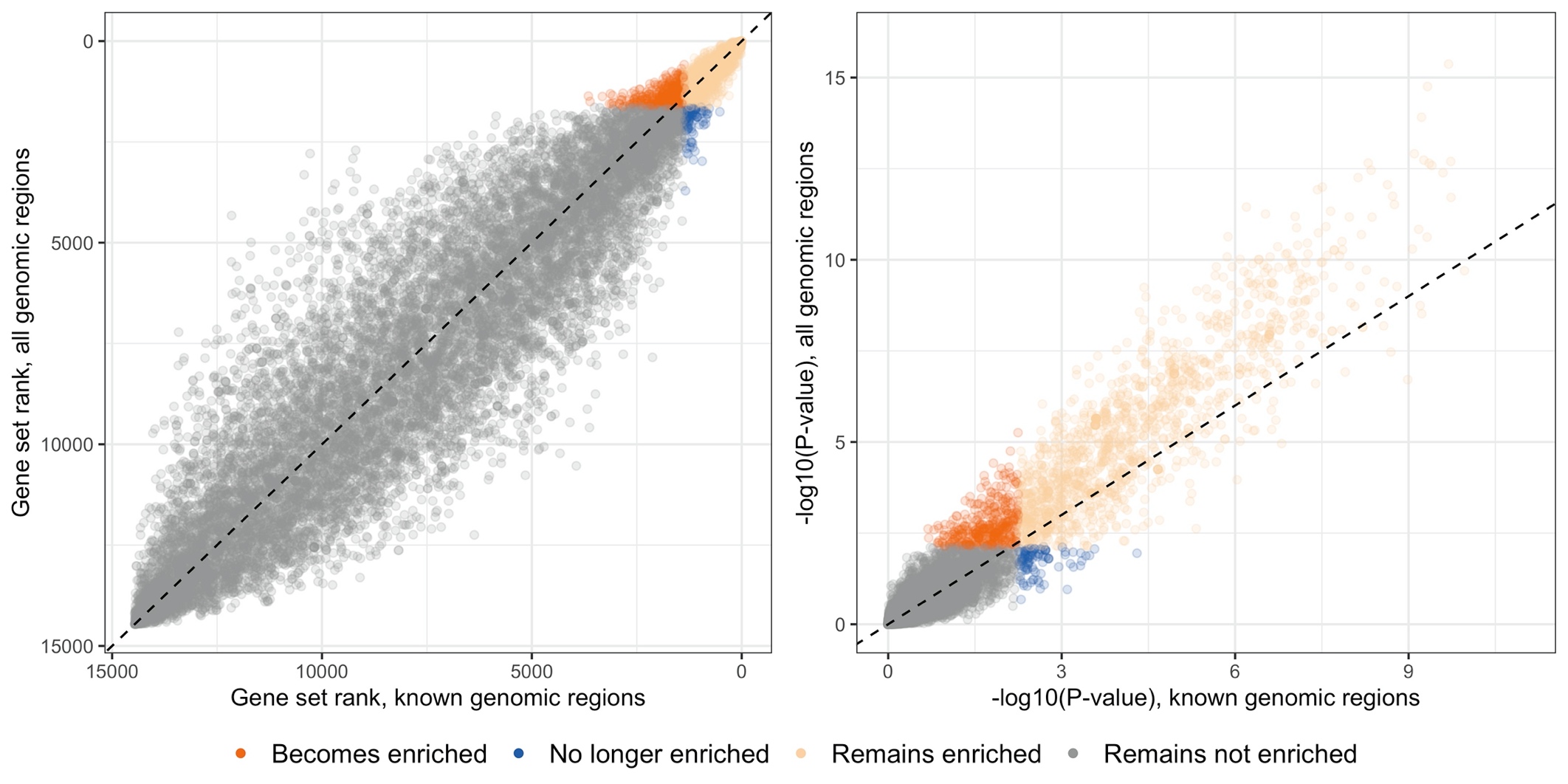

**Supplementary Figure 8 – Functional enrichment of cell-specific TWAS gene clusters**

Genes from each cluster were used to conduct functional enrichment analysis against the Gene Ontology and KEGG databases; y-axis: top significant functions; x-axis: gene clusters; dot size: magnitude of fold change (FC); dot color: significance level, –log_10_(p-value) (neglogp).

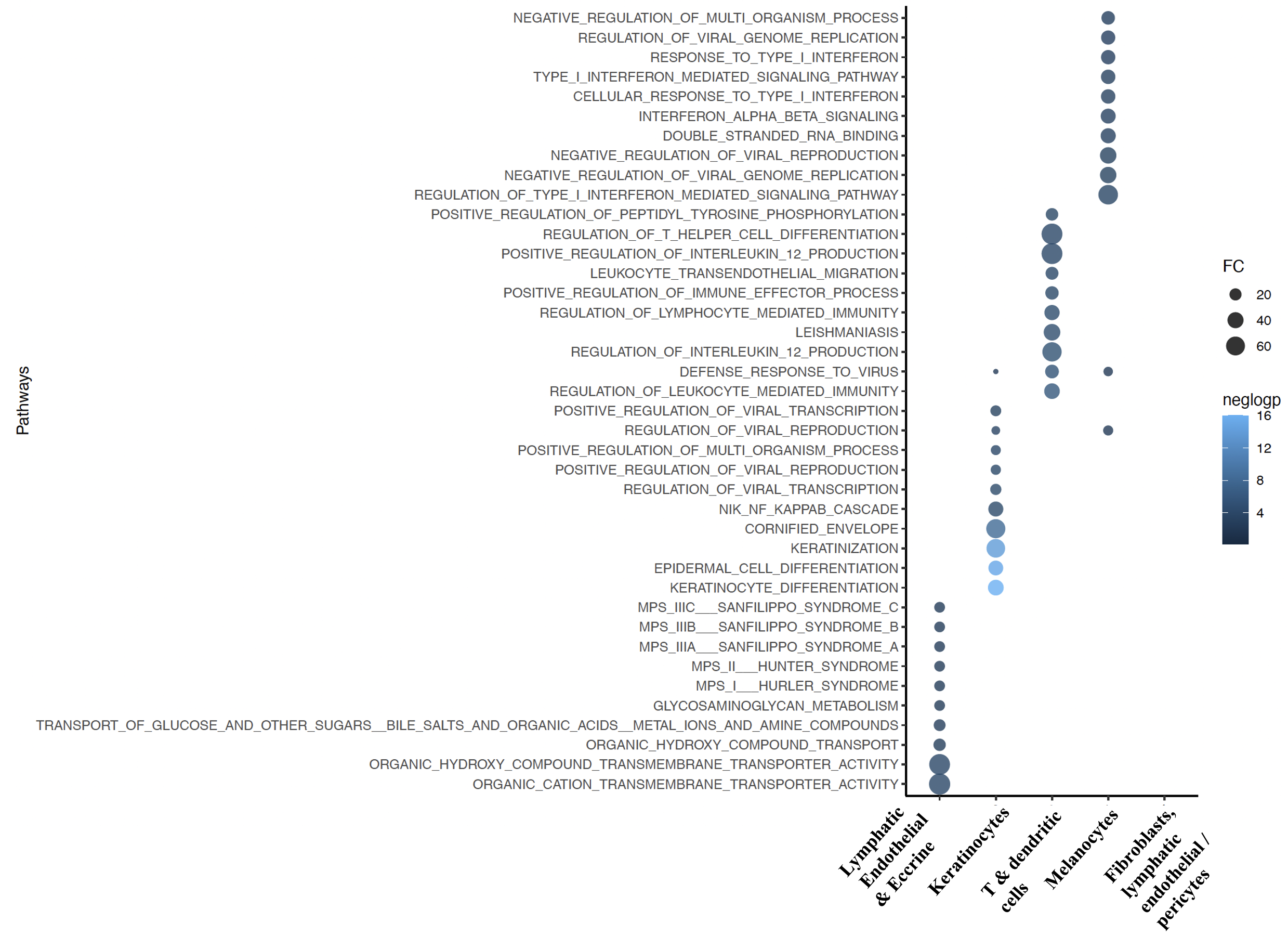

**Supplementary Figure 9 – Enrichment for TWAS genes among cytokine-regulated genes in keratinocytes**

Points indicate cytokine used to stimulate keratinocytes prior to RNA-seq; y-axis: enrichment (observed:expected ratio) for TWAS target genes among the set of genes significantly induced after cytokine stimulation; x-axis: statistical significance of enrichment (measured as -log_10_(FDR)). Coincident points are overplotted, with multiple (bracketed) labels.

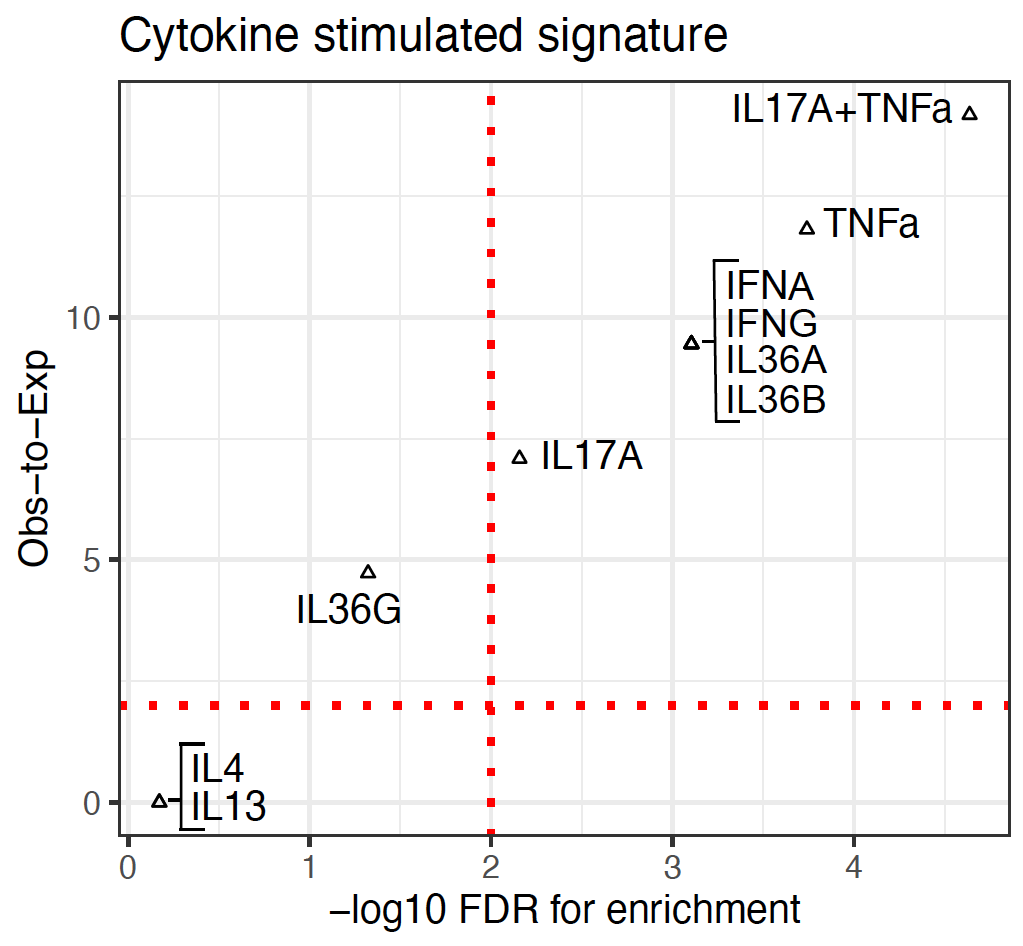

**Supplementary Figure 10 – Genetic correlations with disease and health traits**

Largest estimated genetic correlations with psoriasis. Up to ten traits with statistically significant genetic correlation (P<8.4×10^-5^) are displayed; x-axis, genetic correlation (r_g_); error bars: 95% confidence interval. No negatively correlated disease and health traits reached statistical significance.

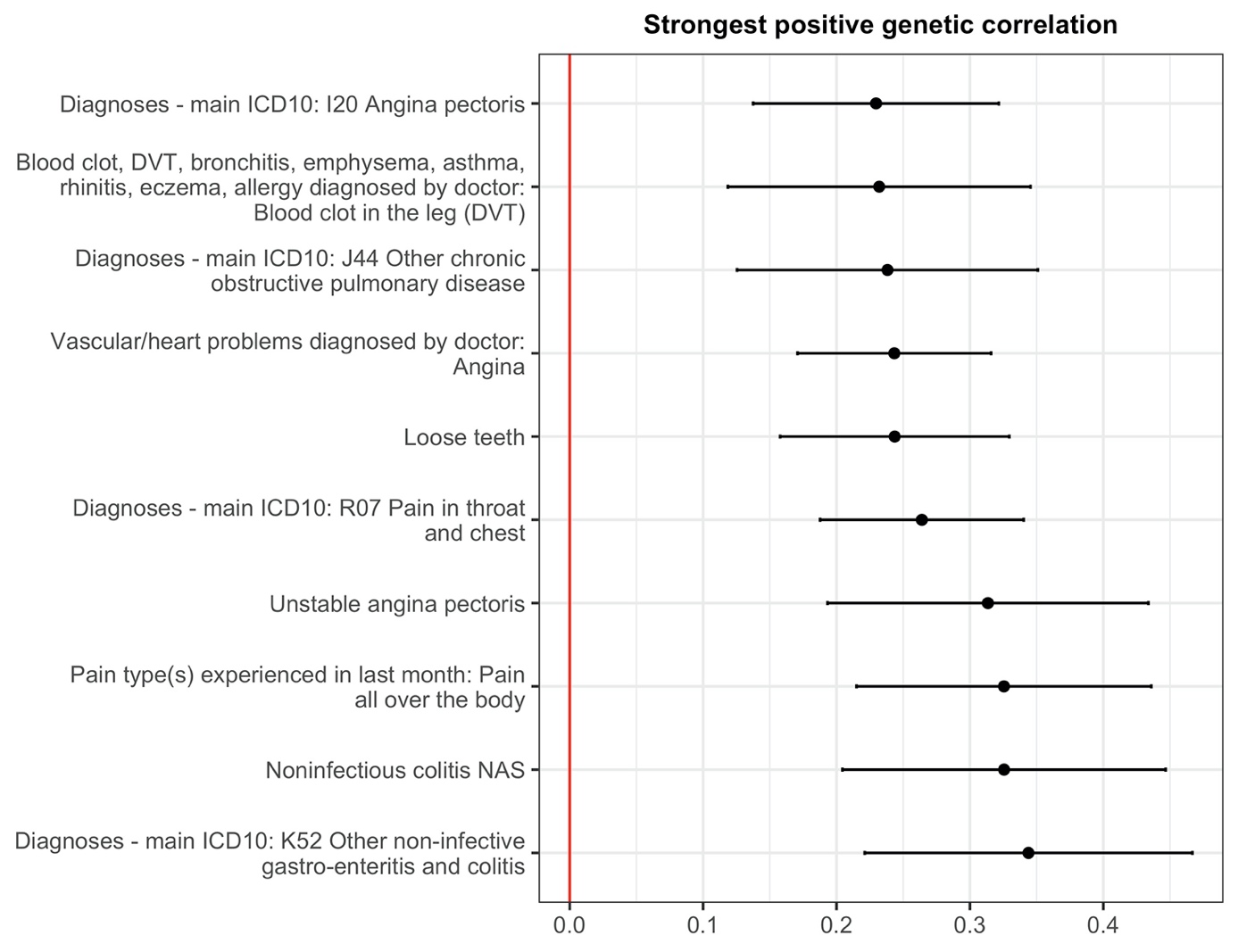

**Supplementary Figure 11 – Genetic correlations with physical, cognitive and biochemical measures**

Largest estimated negative and positive genetic correlations with psoriasis. Up to ten traits with statistically significant genetic correlation (P<8.4×10^-5^) are displayed; x-axis, genetic correlation (r_g_); error bars: 95% confidence interval.

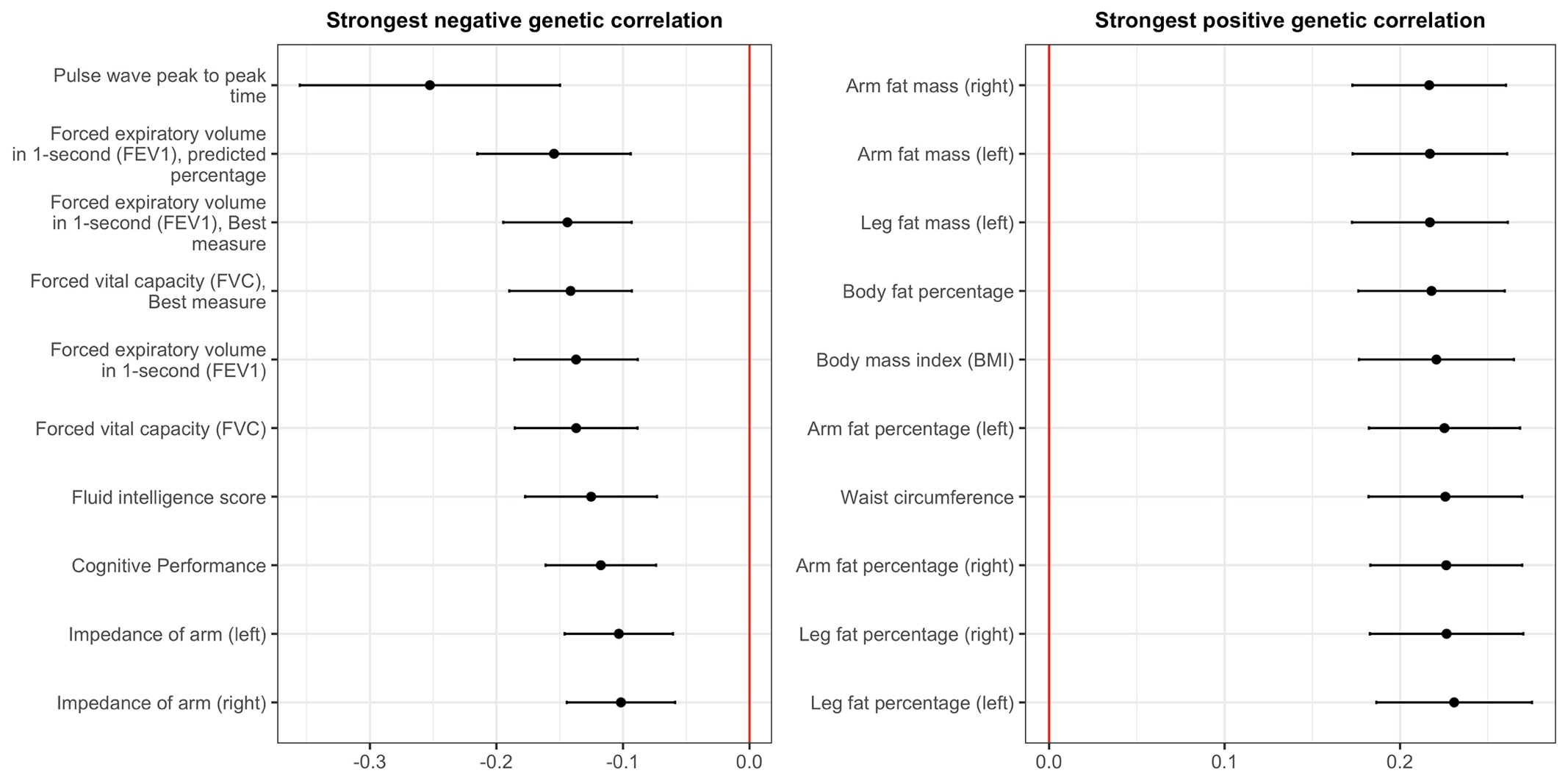

**Supplementary Figure 12 – Genetic correlations with lifestyle and quality of life factors**

Largest estimated negative and positive genetic correlations with psoriasis. Up to ten traits with statistically significant genetic correlation (P<8.4×10^-5^) are displayed; x-axis, genetic correlation (r_g_); error bars: 95% confidence interval.

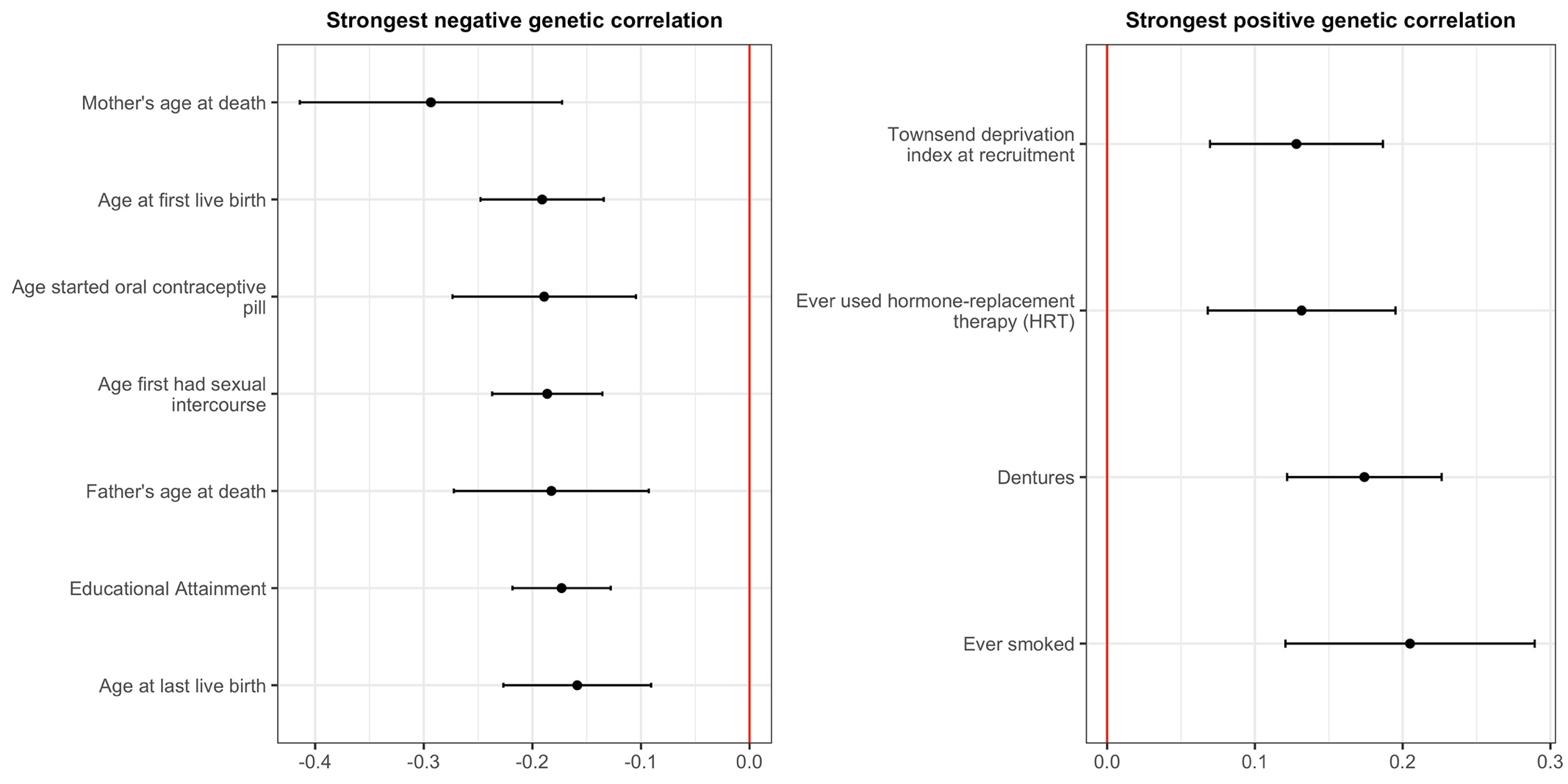

**Supplementary Figure 13 – Latent causal variable analysis**

Estimated causal architecture between psoriasis and other traits. Each point represents a different trait tested; x-axis, genetic causality proportion (GCP), points to the left of the grey dashed line have a causal influence on psoriasis, points to the right are causally influenced by psoriasis; y-axis: statistical significance (z score) for GCP; point colour, genetic correlation (r_g_); point size, magnitude of r_g_; traits with a significant GCP at FDR<0.05 lie above the dashed red line and are highlighted with a grey border. Key to numbering on next page.

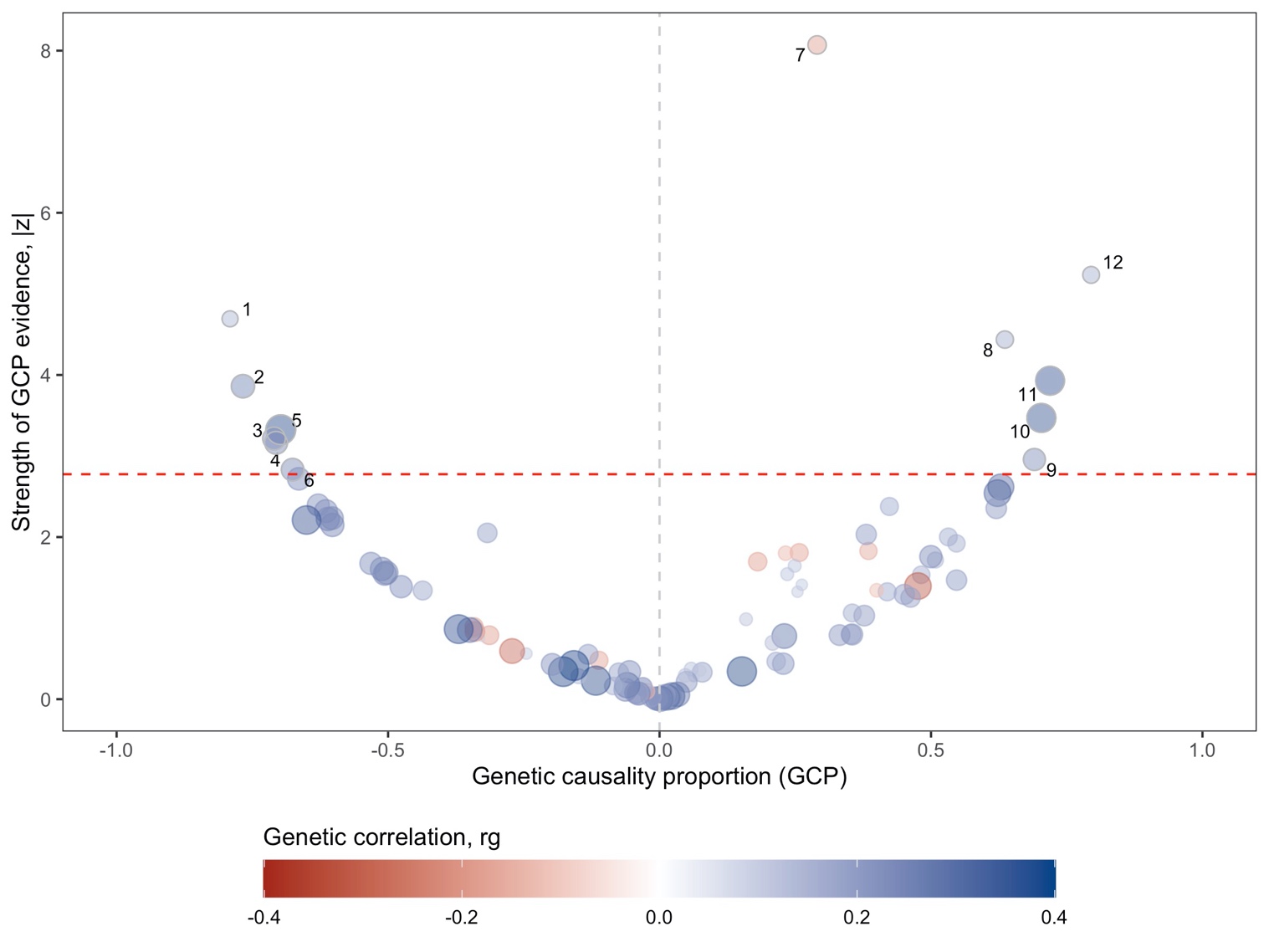

Key for Supplementary Figure 13

| **Negative GCP** (implies trait has causal role in psoriasis) | **Positive GCP** (implies psoriasis has causal role in trait) |
| --- | --- |
| 1. Triglycerides 2. Waist circumference 3. Leg fat mass (right) 4. Leg fat mass (left) 5. Stroke (self-reported) 6. Body mass index (BMI) | 7. Age started oral contraceptive pill 8. Periodontitis + loose teeth 9. Diabetes (self-reported) 10. Fracture at wrist and hand level (hospital in-patient diagnosis) 11. Pain all over the body experienced in last month 12. Back pain experienced in last month |
